# Inferential instability of national sugar and sweetener availability as an indicator of adult obesity trajectories: A global within–between panel audit

**DOI:** 10.64898/2026.08.25.26360957

**Authors:** Zawadi Ally Nkulikwa

**Affiliations:** St John’s University of Tanzania, Dodoma, United Republic of Tanzania

**Keywords:** food balance sheets, ecological inference, fixed effects, temporal diagnostics, inferential stability, cardiometabolic surveillance

## Abstract

National food-balance-sheet data are widely used to characterise population diets, although they measure availability rather than intake. We evaluated whether national sugar and sweetener availability provides an inferentially stable indicator of adult-obesity trajectories. We assembled 3,038 economy–years for 217 economies (2010–2023); the primary complete-case sample comprised 2,153 observations from 160 economies. Exposure was the percentage of dietary-energy supply from FAOSTAT ‘Sugar & Sweeteners’. Within–between and economy-and-year fixed-effects models used economy-clustered standard errors. Temporal specifications were fitted to both their maximal samples and a common sample; a two-part isometric log-ratio analysis addressed composition; and the principal estimator was independently replicated and assessed by wild-cluster bootstrap. A five-percentage-point higher long-run sugar share was associated with 2.37 percentage points higher WHO-modelled adult-obesity prevalence between economies (95% confidence interval 0.52 to 4.23). The adjusted two-way fixed-effects coefficient, reported on the same five-point scale, was −0.68 (−1.18 to −0.19), equivalent to −0.14 points per one-percentage-point higher share. In the common sample (1,228 observations; 156 economies), contemporaneous, one-year-lagged and three-year-lagged coefficients were −0.06 (−0.42 to 0.31), −0.08 (−0.42 to 0.27) and −0.05 (−0.46 to 0.36), respectively; the three-year-future coefficient remained −0.63 (−1.01 to −0.26) and differed from the contemporaneous coefficient by −0.58 (−0.95 to −0.21). The median-equivalent five-point isometric log-ratio estimate attenuated to −0.30 (−0.60 to 0.00; P = 0.051). Independent re-estimation reproduced the principal estimate, and wild-cluster-bootstrap inference supported its computational stability. Economy-specific trends reduced the coefficient to −0.03 (−0.10 to 0.05), and secondary outcomes did not reproduce the obesity pattern. These findings do not indicate that sugar is protective. Across the economies and years examined, national sugar availability did not provide a temporally or construct-stable indicator of change in WHO-modelled adult-obesity prevalence.

## Introduction

### Global cardiometabolic burden and sugar evidence

Obesity has become a defining global health challenge. A pooled analysis of 3,663 population-representative studies estimated that more than one billion children, adolescents and adults were living with obesity in 2022, and the burden increasingly coexists with persistent undernutrition in parts of Africa and South Asia [1]. The World Health Organization (WHO) recommends limiting free sugars because controlled, prospective evidence links higher intake to unhealthy weight gain, dental disease and cardiometabolic risk [2]. Systematic reviews indicate that changes in dietary sugar intake affect body weight largely through energy intake [3], that habitual sugar-sweetened-beverage consumption is associated with type 2 diabetes [4], and that a tightly controlled inpatient trial showed that an ultra-processed dietary pattern can increase ad libitum energy intake and short-term weight gain, even when the diets presented are matched on several nutrients, including sugar [5]. These findings concern individual consumption, products or dietary patterns; they do not establish that a national supply aggregate is an interchangeable measure of what individuals ingest.

The distinction is important because obesity arises within food, labour, social and commercial systems. Obesogenic-environment theory emphasises the conditions that make energy-dense options available, affordable and normal [6]. The commercial-determinants literature extends this focus to corporate practices and market structures [7]. Stress-responsive eating has experimental support in selected populations [8], time scarcity is associated with the use of convenience and ultra-processed foods [9], and long working hours show small, context-dependent associations with weight change [10]. Digital food marketing offers a plausible route via attention, targeting and purchasing friction, but exposure is difficult to measure consistently across economies [11]. Socioeconomic gradients also vary with development, sex and social position rather than following a single universal direction [12]. Accordingly, national averages for inequality, working time or Internet access are structural context indicators, not measures of individual stress, time use or algorithmic advertising exposure.

### The ecological-proxy problem

National food-balance-sheet series are attractive because they are standardised, longitudinal and geographically broad. Earlier cross-national studies reported associations between sugar or soft-drink availability and diabetes and obesity [13,14]. More recent evidence is divided. A 37-country Sub-Saharan African preprint reported a positive between-country relationship that did not persist within countries. In contrast, a Spanish time-series study reported strong predictive performance from Food Balance Sheet nutrient supplies [15,16]. Earlier global longitudinal analyses likewise related national food-supply composition to health outcomes and used panel methods to investigate nutrition-transition questions, but they did not test whether sugar availability retained a stable within-economy interpretation across common-sample temporal, future-exposure and compositional diagnostics [17,18]. These contrasting findings reinforce the need for a globally scoped audit that separates level comparisons from within-economy trajectories. Food balance sheets describe commodities available for human consumption after accounting for production, trade, stocks and non-food uses; they do not capture purchases, household waste, individual intake or the distribution of exposure within a population [19,20]. Validation studies show that concordance with dietary surveys varies by food group and that multi-year averages, energy shares and trends are often more defensible than absolute levels [20–22]. The category ‘Sugar & Sweeteners’ is also broader than free or added sugars and does not capture sugars embedded in every other commodity group.

A second problem is estimand ambiguity. A cross-sectional comparison asks whether economies with higher average sugar availability also have higher average obesity rates. In contrast, a fixed-effects model asks whether departures from an economy’s own expected availability coincide with departures from its own outcome trajectory. These are different questions and may yield different signs. Ecological associations cannot be translated automatically into individual-level effects [23], and fixed effects do not resolve time-varying confounding, differential trends or measurement error. A result is therefore more credible when its direction survives temporal ordering, alternative exposure definitions, weighting and outcome measures, and when a future exposure does not predict an earlier outcome [24]. What remains uncertain is not simply whether national sugar availability correlates with obesity, but whether the measure preserves its direction and interpretation when between-economy contrasts are separated from within-economy change and subjected to temporal and construct-validity tests.

Against this evidence, this study provides a global replication and extension of the reported divergence between cross-sectional and within-country results. It defines a practical boundary of inference: agreement in between-economy rankings does not establish that the same indicator can track within-economy change. Conflating these estimands could mislead surveillance rankings, the interpretation of temporal trends and the evaluation of food-system or policy change. The study evaluates a broader international frame through 2023 and subjects the within-economy association to lag, future-exposure, trend, weighting, exposure-unit and secondary-outcome diagnostics. Its contribution is therefore a diagnostic audit of fitness for a specified ecological use, not criterion validation of individual intake.

### Socio-metabolic systems framework and study objectives

The socio-metabolic systems framework (SMSF) is used only as a secondary organising heuristic. It distinguishes macro-level constructs represented in the panel—structural context, national food availability and modelled cardiometabolic outcomes—from behavioural, commercial and metabolic mechanisms that the data do not measure. Accordingly, Internet penetration cannot represent personalised marketing, and the share working at least 49 hours cannot represent subjective time scarcity or unpaid care. Fig 1 defines the empirical boundary of the study rather than a causal model: the principal contribution is the proxy audit, not validation of individual stress responses, behavioural susceptibility, digital targeting or metabolic pathways.

**Fig 1.**
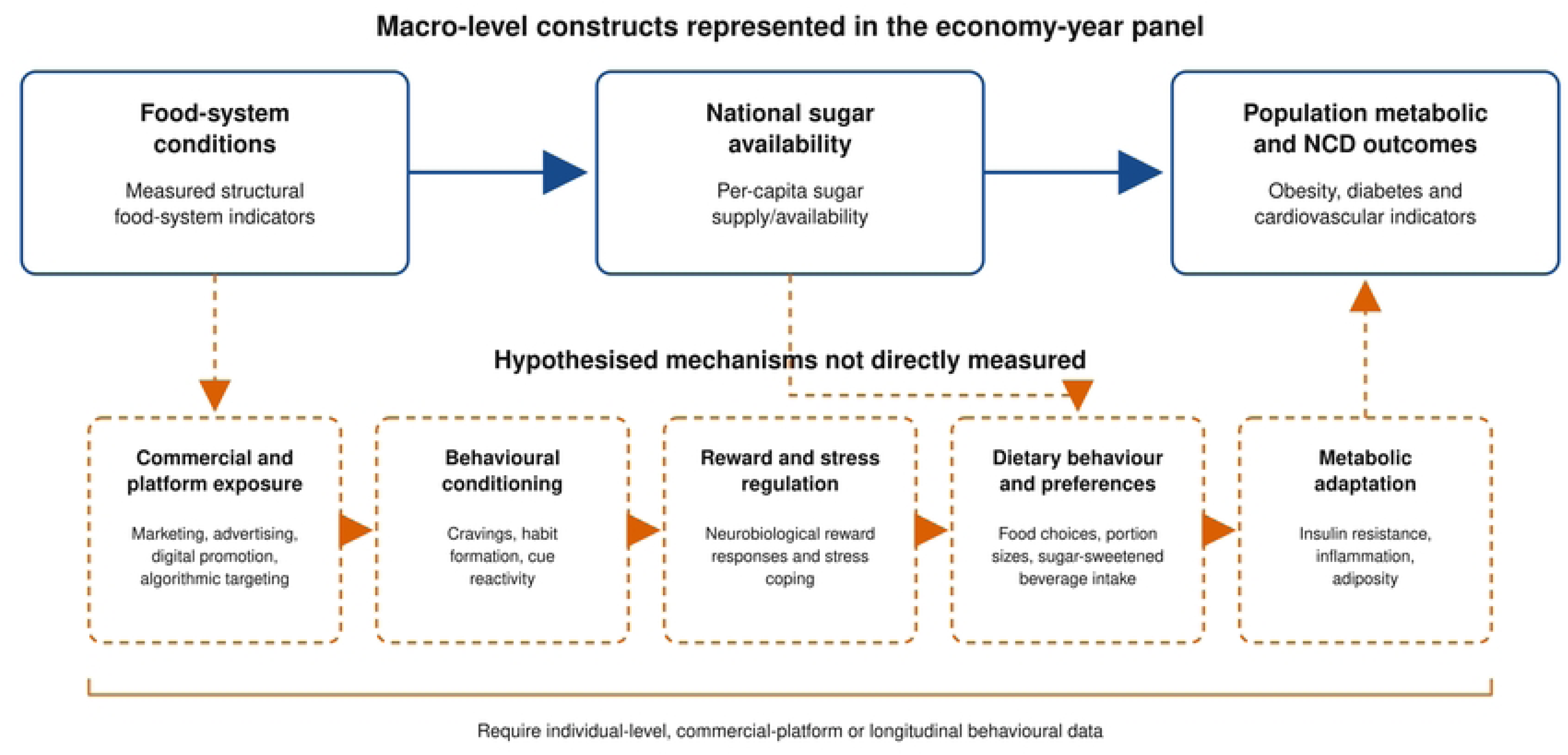
Empirical boundary of the socio-metabolic systems heuristic. Solid elements show macro-level constructs represented in the economy–year panel; dashed elements show hypothesised commercial, behavioural and metabolic mechanisms that require individual-level, platform or longitudinal behavioural data. Arrows indicate conceptual ordering, not identified causal pathways.

Here, inferential stability denotes consistency of an economy-level indicator for the specified use of tracking adult-obesity trajectories. Directional stability required sign consistency among diagnostics addressing the same within-economy use; quantitative stability required broadly comparable coefficients and uncertainty ranges. Between- and within-economy coefficients target different estimands, so their divergence is descriptive context rather than, by itself, proxy failure. Temporal stability was assessed both across model-specific samples and through common-sample coefficients with cluster-aware Wald contrasts. Because no equivalence margin was defined before analysis, non-significant contrasts were not interpreted as proof of equivalence. Statistical significance alone did not define stability.

The primary research question was whether national sugar and sweetener availability provides a directionally and quantitatively stable indicator of within-economy adult-obesity change across temporal specifications, exposure constructions and weighting schemes. Four objectives followed. First, the analysis distinguished between-economy and within-economy associations. Second, it assessed the within-economy estimate using maximal and common temporal samples, a future-exposure temporal diagnostic, economy-specific trends, changes, population weighting, alternative units, a formal compositional balance and a pre-2020 restriction. Third, as secondary exploratory analyses, it examined cross-level modification by disposable-income inequality, long-hours work and digital access. Fourth, it assessed premature non-communicable-disease mortality and raised fasting glucose as secondary outcomes. The study was not preregistered. The three modifier analyses formed a designated multiplicity-adjusted family; the remaining specifications were interpreted as a structured diagnostic audit. Independent estimator replication and wild-cluster-bootstrap inference were reproducibility checks. The directional expectations were positive associations with adverse outcomes and amplification by inequality, time pressure and digital access. A credible longitudinal interpretation was expected to persist under lagged, matched and trend-sensitive models but not to be reproduced by future availability.

## Materials and Methods

### Study design and analytical frame

This ecological longitudinal study compiled an unbalanced economy–year panel for 2010– 2023. The analytical frame comprised 217 World Bank economies and territories with a non-empty geographic-region classification, yielding 3,038 possible economy–years. Regional and income aggregates were excluded. All sources were joined by ISO 3166-1 alpha-3 code and calendar year; FAOSTAT M49 codes were cross-walked to ISO codes. No temporal interpolation or carry-forward was performed. Data and provider metadata were retrieved on 8 August 2026.

The harmonised file incorporated FAOSTAT Food Balances [19,20]; WHO Global Health Observatory (GHO) outcome series disseminated through Our World in Data, with premature NCD mortality additionally disseminated through the World Bank [25–27]; World Development Indicators (WDI) [28]; the Standardised World Income Inequality Database (SWIID) version 9.92 [29,30]; and ILOSTAT working-time series [31]. Adult-obesity and raised-fasting-glucose estimates were provider-attributed WHO GHO series disseminated and minimally processed by Our World in Data for reproducible ISO3–year linkage. Premature NCD mortality was a WHO GHO indicator disseminated via the World Bank and processed by Our World in Data. WHO remained the primary health-data provider. No outcome value was interpolated or re-estimated by the author; other provider values were retained without re-estimation except for transparently derived measures described below.

### Exposure

The primary exposure was the share of national dietary-energy supply represented by FAOSTAT item 2909, ‘Sugar & Sweeteners’. For economy i and year t, the measure was 100 × sugar-and-sweeteners kilocalories per person per day divided by total dietary-energy-supply kilocalories per person per day. Effects were scaled to a five-percentage-point difference. Because this share is compositional, its coefficient in models that also adjust for total dietary energy has an implicit substitution interpretation: at fixed total energy supply, a higher sugar share corresponds to a lower share from other commodities. The formal compositional sensitivity analysis represented the two-part composition as sugar energy and all remaining dietary energy and used the balance z = √(1/2) × ln[sugar kilocalories/(total kilocalories − sugar kilocalories)]. Both components were strictly positive in the analytical sample, so no zero replacement was required. Results were reported per unit of the balance and as the balance change corresponding to an increase from the sample median sugar share (9.789%) to 14.789%. Alternative exposure models used kilograms per person per year (per 10 kg) and kilocalories per person per day (per 100 kcal) as distinct construct checks. These variables estimate national food availability, not free-sugar consumption, household acquisition or individual intake.

### Outcomes

The primary outcome was the WHO age-standardised estimate of adult-obesity prevalence for both sexes aged 18 years or older, defined as a body mass index of at least 30 kg/m² and obtained through the minimally processed WHO GHO series disseminated by Our World in Data [25]. Secondary outcomes were the WHO probability that a person aged 30 years would die before age 70 years from cardiovascular disease, cancer, diabetes or chronic respiratory disease, disseminated through the World Bank and processed by Our World in Data [26], and the WHO age-standardised prevalence of raised fasting blood glucose (at least 7.0 mmol/L) among adults, obtained through the Our World in Data mirror of the WHO GHO series [27]. The obesity series covered 2010–2023; premature NCD mortality covered 2010–2021; and raised fasting glucose covered 2010–2014 in the downloaded files. These were annual modelled national estimates, not independent surveys in every economy–year, and were not relabelled as observed survey values or diagnosed disease.

### Covariates and structural modifiers

The adjustment set was selected to represent major time-varying macro-level common causes, without treating speculative mechanisms as observed. It comprised total dietary-energy supply (per 100 kcal/person/day), log GDP per capita at constant 2021 purchasing-power-parity dollars, urban population share (per 10 percentage points), unemployment (per 5 percentage points), Internet use (per 10 percentage points), population aged 65 years or older (per 5 percentage points), and current health expenditure as a share of GDP. WDI definitions and source agencies were retained [28].

Three cross-level modifiers represent the economic context: the economy-period mean SWIID disposable-income Gini estimate; the economy-period mean share of employed persons working at least 49 hours per week, derived from identical ILOSTAT source records; and the economy-period mean share of individuals using the Internet. Each mean was standardised across economies. SWIID values are harmonised, model-based estimates with attached uncertainty [29,30]; ILOSTAT comparability depends on national source design [31]; and Internet use measures connectivity rather than marketing exposure. Interactions therefore test contextual patterning, not individual susceptibility.

### Statistical estimands

We first summarised total, between-economy and within-economy dispersion. Population-weighted annual means, unweighted economy means and long-period changes were presented descriptively. Spearman coefficients in the descriptive analysis captured rank association and were not used as causal estimates.

A correlated-random-effects decomposition [32] entered the economy mean and economy-mean-centred deviations for sugar and each covariate, with year and region indicators. The between coefficient compares economies with different long-run average availability; the within coefficient compares deviations from an economy’s long-run mean. The principal longitudinal specification was ordinary least squares with economy and year fixed effects:

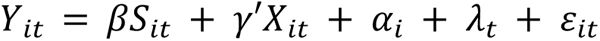

where Y is the outcome prevalence or probability in percentage points, S is the sugar energy share divided by five, X is the adjustment vector, α is an economy fixed effect, and λ is a year fixed effect. The coefficient β is an associational within-economy contrast, not a causal effect. Standard errors used the CR1 small-sample correction and were clustered by economy [33]; a sensitivity model was clustered by economy and year. Statistical tests were two-sided. Pairwise differences among common-sample temporal coefficients were evaluated with Wald tests using a stacked cross-equation, economy-clustered covariance. For the three designated secondary exploratory modifier tests, statistical evidence was evaluated at α = 0.05 after Benjamini–Hochberg correction.

### Model assumptions and uncertainty

The fixed-effects coefficient was estimated from within-economy exposure changes after adjustment for common year shocks, the specified time-varying covariates and all time-invariant economy characteristics. Its interpretation additionally requires an approximately linear and additive conditional association over the observed range, adequate temporal alignment, and no consequential omitted time-varying common cause, differential measurement error or anticipatory process. These are identifying assumptions rather than empirical facts established by the model. Cross-level interactions were limited to the three stated contextual modifiers; the analysis does not claim to exhaust plausible non-linearity or effect heterogeneity.

Provider point estimates were analysed as fixed inputs. The regression confidence intervals quantify uncertainty conditional on those inputs and the stated covariance estimator; they do not propagate the estimation uncertainty embedded in WHO modelled outcomes, SWIID imputations or other harmonised indicators. Consequently, the intervals may understate total data-generating uncertainty and the coefficients should be interpreted as associations among constructed population indicators rather than direct measurements.

### Temporal, construct and sample diagnostics

Temporal diagnostics used exact one- and three-year exposure lags and a three-year future-exposure lead. The lead was interpreted as a temporal diagnostic rather than a definitive negative control: a future value cannot cause an earlier outcome, but a persistent exposure can remain correlated with earlier exposure and omitted processes spanning time [24]. Each specification was first fitted to its maximal eligible sample. To isolate temporal alignment from changing sample composition, we then required the outcome at year t, contemporaneous, one-year-lagged, three-year-lagged and three-year-future exposures, and every covariate to be simultaneously observed. The resulting common sample comprised 1,228 observations from 156 economies during 2013–2020. Four otherwise identical economy-and-year fixed-effects models were fitted to those rows. Pairwise coefficient contrasts used a cross-equation covariance constructed from stacked economy-level score vectors with the same CR1 correction. Additional specifications included economy-specific linear time trends, annual first differences, five-year differences, population-weighted fixed effects and a restriction to 2010–2019. Construct sensitivity included the formal ILR balance and replaced the energy share with kilograms and absolute kilocalories. Sample checks retained economies with at least ten complete years and excluded populations below one million and observations above the 99th exposure percentile. Full definitions and contrasts are reported in S3 Appendix and S5 Results.

Dynamic-panel estimators were not used because conditioning on lagged obesity would alter the estimand and, in a short, highly persistent panel, would require additional assumptions not warranted by this diagnostic, non-causal design. The high residual AR(1) was therefore treated as a limitation and an identification warning rather than as a defect that a more complex estimator could automatically remedy.

Cross-level models interacted the within-economy sugar measure with each standardised economy-period contextual mean, retaining the primary controls and the time-varying value of the modifier where applicable. False-discovery-rate q values were calculated across the three secondary exploratory interaction tests using the Benjamini–Hochberg procedure [34]. Secondary outcomes used the primary contemporaneous and three-year-lag specifications.

The study was not preregistered. The three secondary exploratory modifier tests formed the only designated multiplicity-adjusted hypothesis family. The remaining specifications were interpreted jointly as a structured coherence and diagnostic audit of the primary estimand, rather than as independent confirmatory hypothesis tests; inference rests on concordance of direction, magnitude and diagnostic behaviour rather than on selecting isolated P values.

### Missing data, software and reproducibility

Analyses were outcome-specific complete cases. Missing values were not imputed because absence often reflected source coverage, survey timing or modelled-series availability rather than an exchangeable random process. The primary adjusted sample contained 2,153 economy–years from 160 economies (mean 13.5 observations per economy), and the common temporal sample contained 1,228 observations from 156 economies. Model N, economy count and year range are reported for every estimate. Supplementary Table S1 in S3 Appendix documents the cascade from the 3,038 economy–year frame through exposure, outcome and model-specific complete cases. No missing-at-random claim is made: complete-case estimates are conditional on inclusion in the observed analytical panel, and selection related to statistical capacity or development may limit transportability.

Data processing and estimation were implemented in Python 3.12.13 using pandas 2.2.3, NumPy 2.3.5, SciPy 1.17.0, scikit-learn 1.8.0, Matplotlib 3.10.8 and Pillow 12.2.0. The principal ordinary-least-squares and CR1 covariance estimates were implemented directly from matrix expressions. Independently, scikit-learn LinearRegression re-estimated the full-dummy economy-and-year fixed-effects model, after which CR1 covariance was reconstructed from independently aggregated economy score vectors. Coefficient and standard-error agreement was required within 1 × 10⁻¹⁰. A restricted Rademacher wild-cluster-bootstrap-t test used 999 repetitions and seed 20260811 [38]. Duplicate ISO-year keys, numeric ranges, exact calendar lags, formula-derived fields, source updates and row-level completeness flags were checked before modelling. The clean run exports every coefficient, contrast, diagnostic, metadata snapshot and figure, together with SHA-256 checksums. TIFF figures are flattened RGB files at 350 dpi with LZW compression. Reporting was guided by STROBE recommendations adapted to the ecological longitudinal design; the completed checklist is S4 Checklist.

### Artificial intelligence tools and technologies

OpenAI ChatGPT Work (Codex, GPT-5 and GPT-5.6; accessed 9–12 August 2026) supported statistical coding, figure generation, structural organisation, drafting and substantial language editing across the manuscript and supporting materials. The author checked merge keys and source metadata against official provider files, reran the complete analysis, reviewed model diagnostics and visual outputs, and verified references, numerical statements and substantive interpretations against the cited sources and reproducible outputs. No confidential, proprietary or individual-level human-participant data were processed; all analysed data were public, aggregated economy–year statistics. The author reviewed and approved the final text and accepts full responsibility for the study’s integrity, accuracy and conclusions.

### Ethics

The study used only publicly available, aggregated economy–year statistics and contained no individual, identifiable or confidential information. Human-participant consent and institutional ethics review were therefore not required.

## Results

### Coverage and descriptive patterns

Sugar availability was observed in 2,397 economy–years across 177 economies; adult obesity was observed in 2,744 economy–years across 196 economies. The primary adjusted sample represented 70.9% of the full frame and contained 2,153 observations from 160 economies. The common temporal sample contained 1,228 observations from 156 economies during 2013–2020. All World Bank regions were represented in the primary model: Europe and Central Asia contributed 662 observations from 48 economies; sub-Saharan Africa 501 from 36; Latin America and the Caribbean 375 from 27; the Middle East and North Africa 263 from 21; East Asia and the Pacific 249 from 20; South Asia 75 from six; and North America 28 from two. Coverage was lower for SWIID inequality (1,727 observations in the interaction model), working time (1,248), premature NCD mortality (1,853) and raised fasting glucose (776).

The mean sugar and sweeteners supply was 9.94% of dietary energy (total SD 4.19; between-economy SD 4.07; within-economy SD 1.09). Thus, most variation in exposure was cross-sectional rather than temporal. The mean adult obesity prevalence was 21.06% (between-economy SD 12.69; within-economy SD 1.88). The population-weighted sugar energy share rose modestly from 8.12% in 2010 to 8.35% in 2016, then fell to 7.56% in 2023; the population-weighted adult obesity increased monotonically from 10.98% to 15.64%. These simultaneous but differently shaped trends make a naïve contemporaneous fixed-effects coefficient vulnerable to trend structure. Table 1 summarises coverage and dispersion of the core variables.

**Table 1.** Economy–year coverage and dispersion of core variables, 2010–2023.

| Construct | Obs.<br>(econ.) | Years | Mean<br>(SD) | Btw.<br>SD | Within<br>SD | Range |
| --- | --- | --- | --- | --- | --- | --- |
| Sugar energy share (%) | 2,397<br>(177) | 2010–<br>2023 | 9.94<br>(4.19) | 4.07 | 1.09 | 0.70–<br>21.90 |
| Sugar supply (kg/person/year) | 2,397<br>(177) | 2010–<br>2023 | 41.47<br>(25.38) | 24.53 | 6.57 | 2.91–<br>179.22 |
| Adult obesity (%) | 2,744<br>(196) | 2010–<br>2023 | 21.06<br>(12.80) | 12.69 | 1.88 | 0.70–<br>75.61 |
| Premature NCD mortality (%) | 2,220<br>(185) | 2010–<br>2021 | 20.01<br>(7.26) | 7.18 | 1.20 | 6.90–<br>44.10 |
| GDP per capita, PPP (\$000) | 2,781<br>(199) | 2010–<br>2023 | 25.24<br>(25.95) | 25.67 | 4.10 | 0.99–<br>174.57 |
| Disposable-income Gini (points) | 2,046<br>(181) | 2010–<br>2023 | 37.78<br>(7.65) | 7.76 | 0.91 | 22.30–<br>64.80 |
| Working $\geq 49$ hours (%) | 1,349<br>(168) | 2010–<br>2023 | 17.06<br>(11.81) | 12.27 | 3.87 | 0.47–<br>97.00 |
| Urban population (%) | 3,038<br>(217) | 2010–<br>2023 | 60.82<br>(23.76) | 23.74 | 1.92 | 10.98–<br>100.00 |
| Unemployment (%) | 2,614<br>(187) | 2010–<br>2023 | 7.89<br>(5.97) | 5.71 | 1.82 | 0.10–<br>36.47 |
| Internet users (%) | 2,712<br>(209) | 2010–<br>2023 | 52.62<br>(30.12) | 27.10 | 13.67 | 0.25–<br>100.00 |
| Population aged $\geq 65$ (%) | 3,038<br>(217) | 2010–<br>2023 | 8.96<br>(6.36) | 6.28 | 1.10 | 0.86–<br>37.32 |
| Health expenditure (% GDP) | 2,678<br>(193) | 2010–<br>2023 | 6.52<br>(2.96) | 2.78 | 1.03 | 1.33–<br>27.09 |

Notes: Units are stated in the construct labels; GDP is expressed in thousands of constant 2021 international dollars at purchasing-power parity. Obs. (econ.), non-missing economy–years (economies); SD in the mean column is the total SD. Between SD is the standard deviation of economy means. Within SD is calculated from deviations around each economy mean. Statistics use all available observations for each variable and are not restricted to the primary complete-case sample.

The global trends, between-economy levels and within-economy changes are displayed in Fig 2.

**Fig 2.**
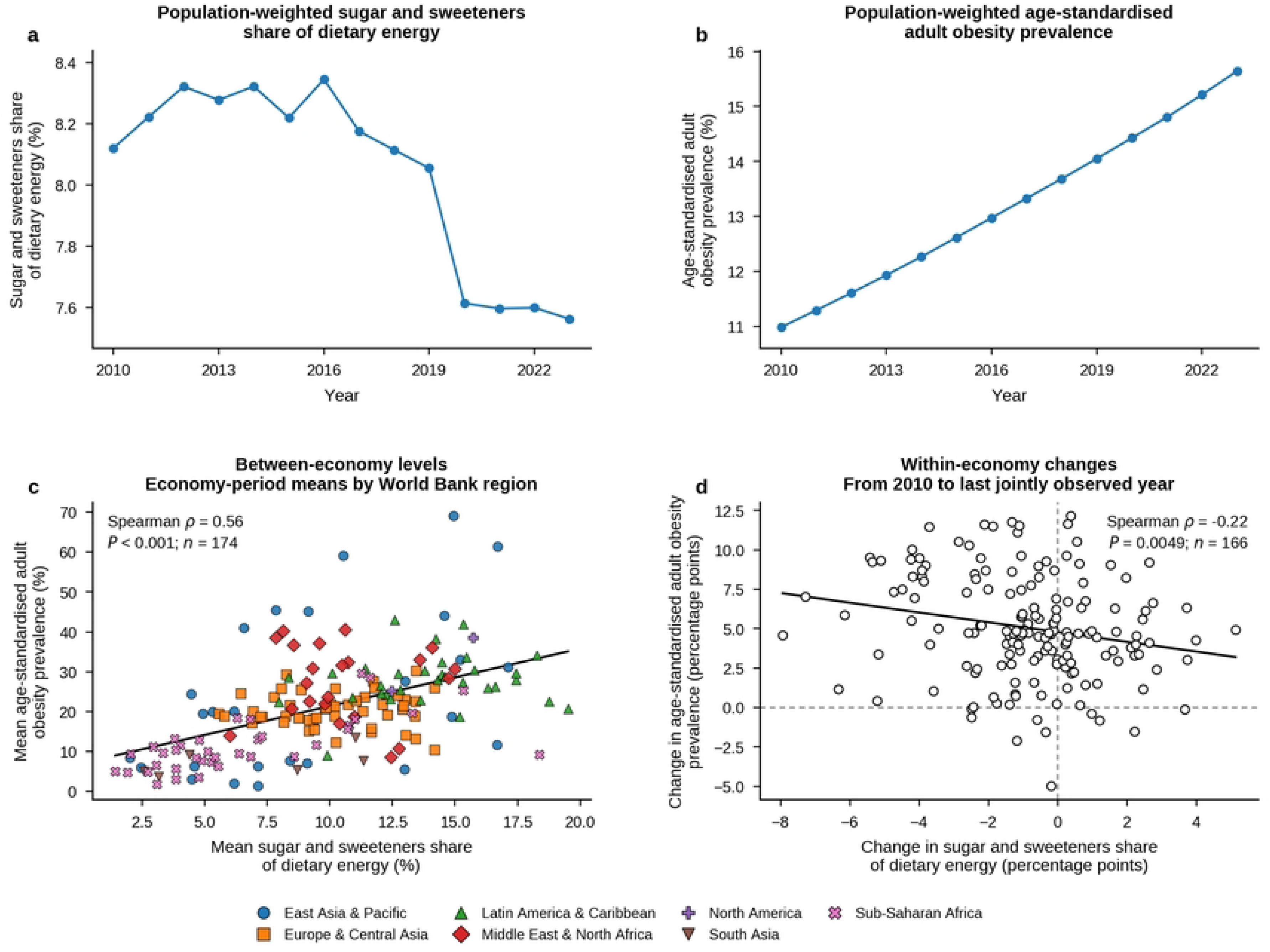
Descriptive separation of global trends, between-economy levels and within-economy changes. a, Population-weighted sugar and sweetener share of dietary energy. b, Population-weighted WHO-modelled age-standardised adult-obesity prevalence. c, Economy-period means by World Bank region (n = 174 economies). d, Change from 2010 to each economy’s last jointly observed year among economies with joint exposure–outcome observations in 2010 (n = 166). Because endpoints vary across economies in panel d, that panel is an exploratory unequal-duration comparison rather than a common-period change estimate. Lines in a and b connect annual population-weighted estimates; lines in c and d are descriptive linear fits. Spearman coefficients are unadjusted rank associations and do not identify causal effects.

### Between-economy and within-economy estimates

The adjusted within–between decomposition produced sharply different estimands. A 5-percentage-point higher long-run sugar energy share was associated with 2.37 percentage points higher WHO-modelled adult-obesity prevalence between economies (95% CI 0.52 to 4.23; P = 0.013). The within-economy deviation was −0.58 points and compatible with both a modest inverse and a null association (95% CI −1.61 to 0.45; P = 0.269). Because the 5-point reporting contrast is substantially larger than the observed within-economy SD of 1.09 points, it provides a common comparison scale rather than representing a typical annual change. This contrast mirrors Fig 2: economies with higher mean sugar shares tended to have higher mean obesity (Spearman ρ = 0.56), whereas long-period changes were negatively ranked (ρ = −0.22). Magnitudes and confidence intervals, rather than thresholded P values alone, were used to judge coherence across specifications.

The exposure-only two-way fixed-effects (TWFE) model estimated −0.64 percentage points per 5-point higher sugar share (95% CI −1.15 to −0.14; S3 Table S5). Adjustment produced −0.68 (−1.18 to −0.19; P = 0.007). On smaller reporting scales, this equals −0.14 percentage points per 1-percentage-point higher share and approximately −0.15 points per one within-economy SD of exposure (1.09 points). The one-year-lag estimate was −0.54 (−1.01 to −0.07), while the three-year lag attenuated to −0.21 (−0.64 to 0.22). These coefficients are associational departures around economy and year means; their inverse sign does not by itself indicate benefit. Table 2 reports the principal adult-obesity estimates, with additional diagnostics in S3 Table S5.

**Table 2.**
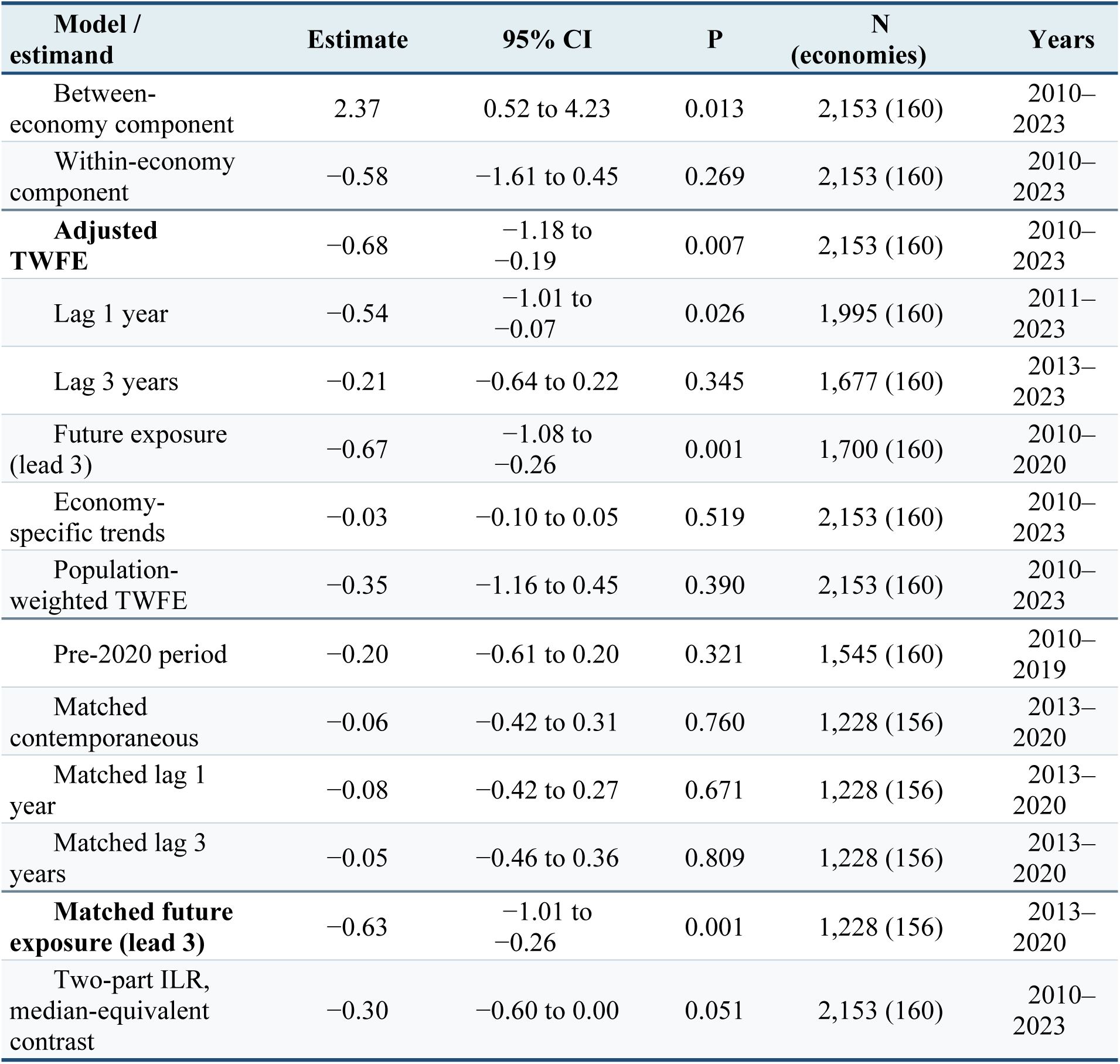
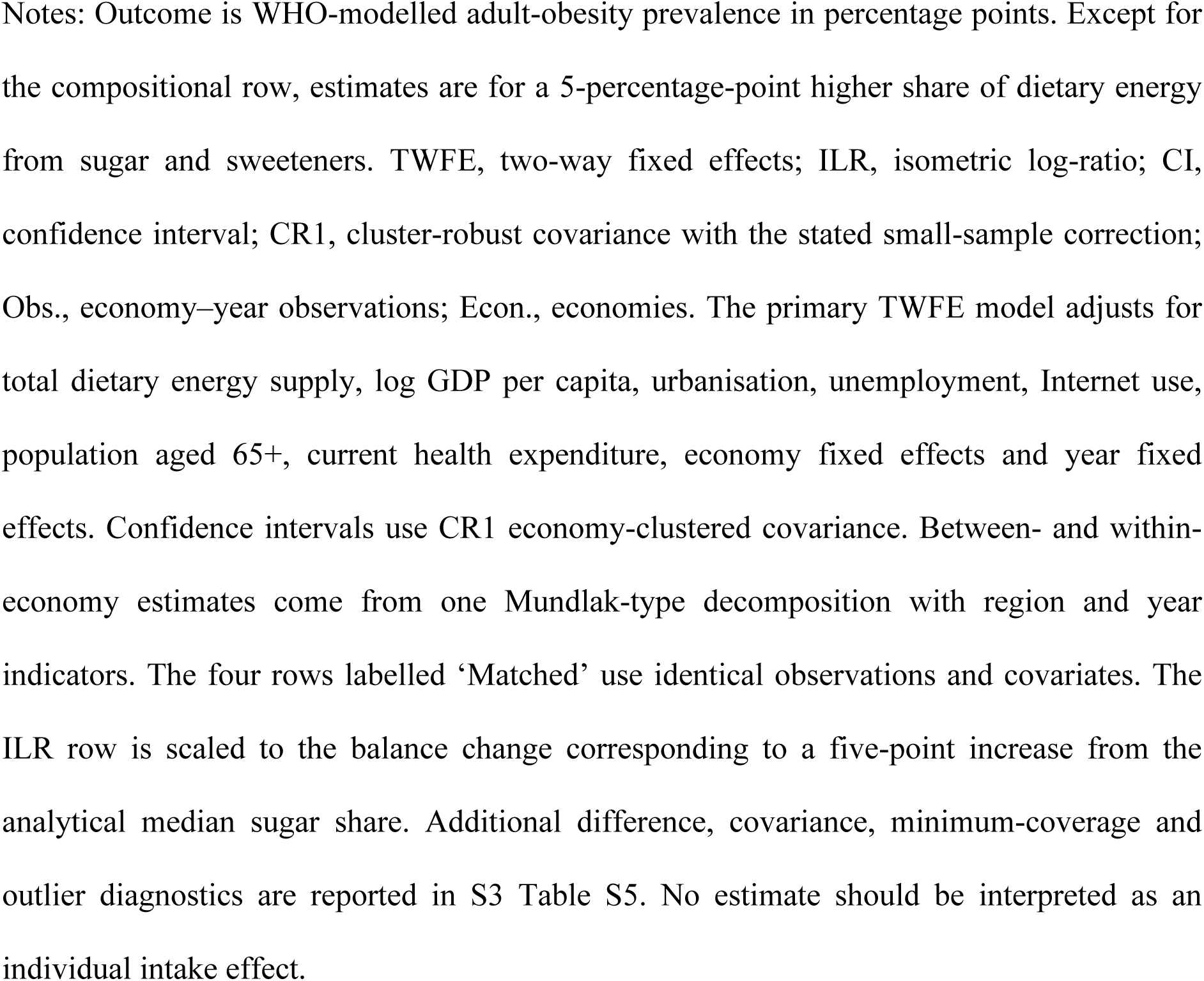
Within–between and principal diagnostic estimates for WHO-modelled adult-obesity prevalence.

### Temporal diagnostics and construct sensitivity

The future-exposure diagnostic weakened a causal interpretation of the contemporaneous inverse association. In the original model-specific samples, sugar availability three years after the outcome year was associated with current obesity at −0.67 percentage points (95% CI −1.08 to −0.26; P = 0.001), whereas the three-year-lag estimate was −0.21 (−0.64 to 0.22). In the common sample of 1,228 observations from 156 economies, the contemporaneous, one-year-lagged and three-year-lagged coefficients were −0.06 (−0.42 to 0.31; P = 0.760), −0.08 (−0.42 to 0.27; P = 0.671) and −0.05 (−0.46 to 0.36; P = 0.809), respectively. The matched future-exposure coefficient remained −0.63 (−1.01 to −0.26; P = 0.001). Its difference from the contemporaneous coefficient was −0.58 (−0.95 to −0.21; Wald P = 0.002), and its difference from the one-year lag was −0.56 (−1.05 to −0.07; P = 0.025); the difference from the three-year lag was −0.58 (−1.19 to 0.03; P = 0.060). The other matched contrasts were near zero (all P ≥ 0.876; Supplementary Table S2). Matching therefore removed the original contemporaneous and lagged inverse coefficients but not the future-exposure association, strengthening the temporal-instability diagnosis rather than attributing it solely to changing sample composition.

The formal two-part ILR model estimated −0.91 percentage points per unit increase in √(1/2) × ln(sugar energy/non-sugar energy) (95% CI −1.82 to 0.003; P = 0.051). At the analytical median, the balance change equivalent to increasing the sugar share from 9.789% to 14.789% corresponded to −0.30 points (−0.60 to 0.001; P = 0.051). The direction remained inverse, but the magnitude attenuated and the interval included the null; the conventional 0.05 inference from the primary ratio model therefore did not survive formal compositional treatment. Annual and five-year change models remained inverse (−0.08 and −0.46 points), but economy-specific trends reduced the coefficient to −0.03 (−0.10 to 0.05), the pre-2020 estimate was −0.20 (−0.61 to 0.20), and population weighting yielded −0.35 (−1.16 to 0.45).

Estimates per 10 kg and per 100 kcal were −0.02 (−0.21 to 0.17) and −0.34 (−0.70 to 0.02), respectively. The independent full-dummy implementation reproduced the principal coefficient and CR1 standard error within 1 × 10⁻¹⁰; the 999-repetition wild-cluster-bootstrap-t P value was 0.010. Fig 3 summarises the temporal, matched, compositional, sample and outcome sensitivity estimates.

**Fig 3.**
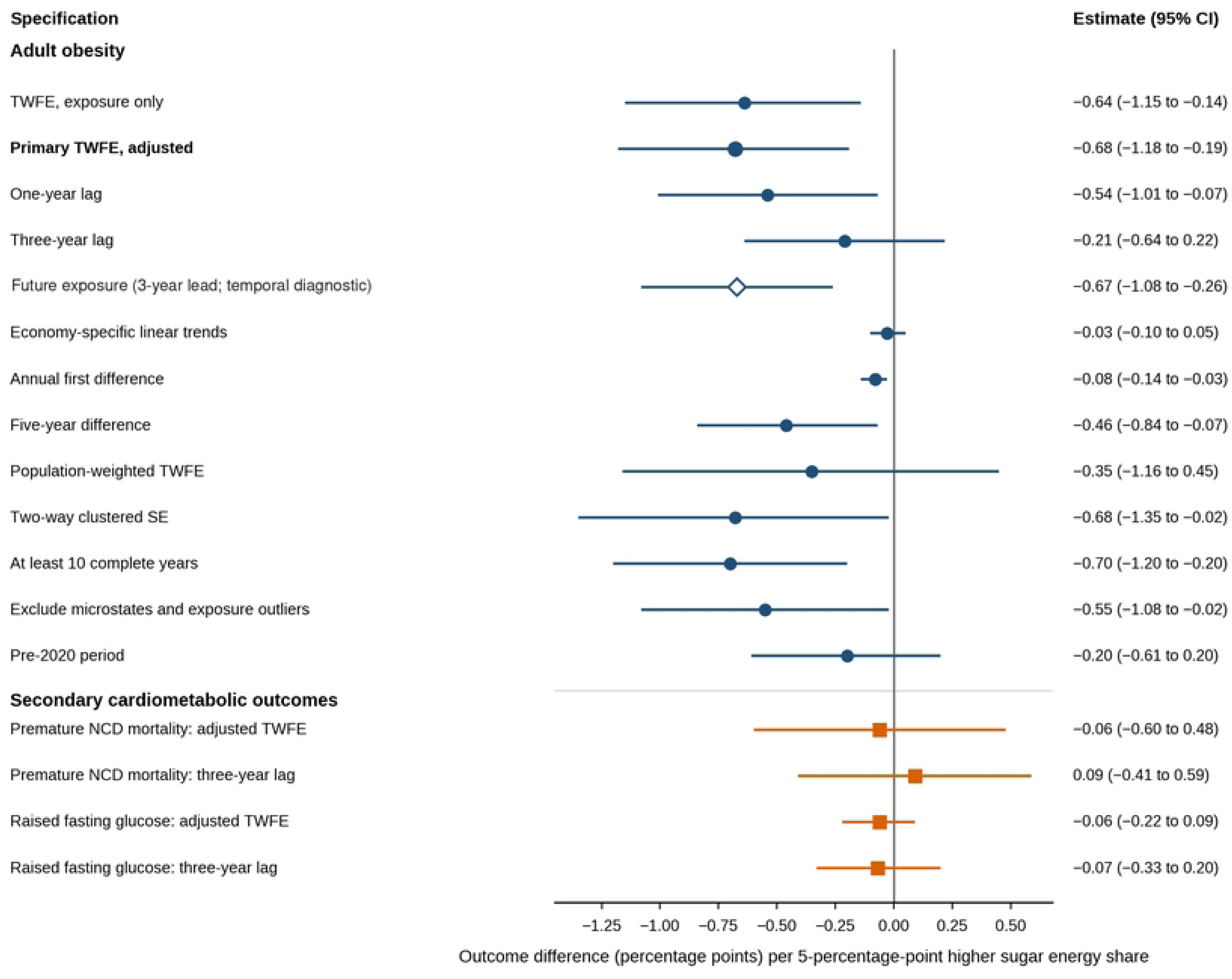
Association estimates across primary, temporal, compositional and outcome specifications. Points show percentage-point outcome differences per comparable five-point exposure contrast; horizontal lines indicate 95% confidence intervals. Blue rows use the original model-specific samples, teal rows use one common temporal sample (1,228 observations; 156 economies; 2013–2020), purple shows the two-part ILR balance scaled from the median sugar share, and orange shows secondary outcomes. Open diamonds identify future-exposure temporal diagnostics. Raised fasting glucose is available only through 2014. All estimates are associational.

### Secondary exploratory structural modification

In the secondary exploratory analyses, disposable-income inequality shifted the unstable inverse fixed-effects association further in a negative direction, opposite to the directional expectation. Each one-SD higher economy-period mean Gini (7.76 points) changed the sugar coefficient by −0.66 percentage points (95% CI −1.12 to −0.20; P = 0.005; false-discovery-rate q = 0.015). This interaction is not evidence that inequality is protective or that sugar is beneficial. The long-hours interaction was −0.11 (−0.74 to 0.52; P = 0.737), and the digital-access interaction was 0.15 (−0.32 to 0.63; P = 0.518); both intervals encompassed substantively different directions. Table 3 reports the designated multiplicity-adjusted family.

**Table 3.** Secondary exploratory cross-level modification of the within-economy sugar– obesity association.

| Context | Mean (SD) | $\Delta\beta$ | 95% CI | P; q | N (economies) |
| --- | --- | --- | --- | --- | --- |
| Income inequality | 38.58 (7.76) | $-0.66$ | $-1.12$ to $-0.20$ | $0.005$; $0.015$ | 1,727 (150) |
| Long-hours work | 19.31 (12.27) | $-0.11$ | $-0.74$ to $0.52$ | $0.737$; $0.737$ | 1,248 (142) |
| Digital access | 52.47 (27.10) | $0.15$ | $-0.32$ to $0.63$ | $0.518$; $0.737$ | 2,153 (160) |

Notes: $\Delta\beta$ is the change in the adult-obesity coefficient associated with a one-standard-deviation increase in the economy-period mean context. SD, standard deviation; CI, confidence interval; Obs., economy–year observations; econ., economies; q, Benjamini–Hochberg false-discovery-rate-adjusted P value; SWIID, Standardised World Income Inequality Database. Income inequality is measured by the SWIID disposable-income Gini. Sugar exposure remains scaled per 5 percentage points. These analyses were secondary and exploratory, were not preregistered, and formed one designated multiplicity-adjusted family. Context means are structural proxies and do not measure individual susceptibility.

The secondary exploratory interaction estimates and their confidence intervals are displayed in Fig 4.

**Fig 4.**
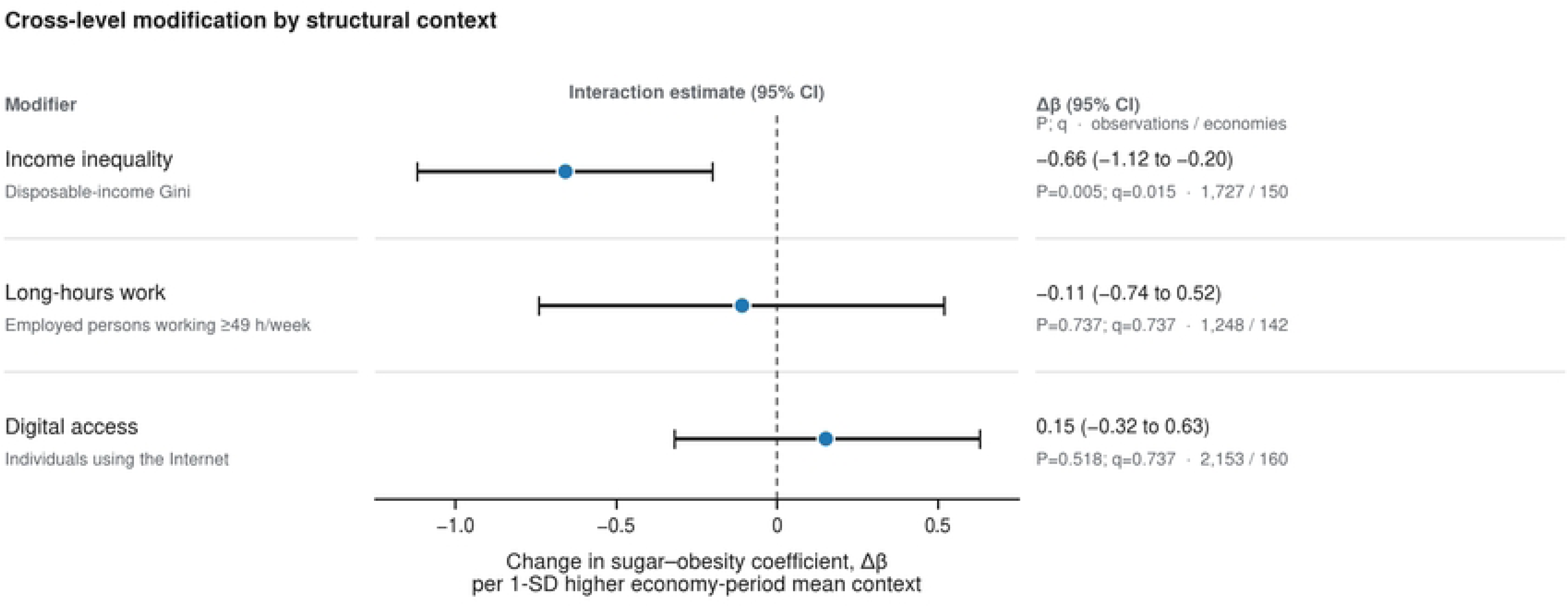
Secondary exploratory cross-level modification by structural context. Points show the estimated change in the sugar–obesity coefficient per 1-SD increase in each economy-period mean contextual modifier; horizontal lines indicate 95% confidence intervals. The income-inequality interaction is opposite to the directional expectation. Long-hours work and Internet use are coarse structural proxies and should not be interpreted as direct measures of time scarcity or algorithmic food-marketing exposure.

### Secondary outcomes

For premature NCD mortality, the contemporaneous estimate was near zero (−0.06 percentage points; 95% CI −0.60 to 0.48; P = 0.835), and the three-year-lag estimate was 0.09 (−0.41 to 0.59; P = 0.724). The raised-fasting-glucose estimate was also near zero contemporaneously (−0.06; −0.22 to 0.09; P = 0.427). Each interval remained compatible with inverse and positive associations of modest magnitude. The lagged raised-glucose estimate was imprecise and based solely on 2013–2014 outcomes; it is reported for completeness rather than substantive longitudinal interpretation. The negative contemporaneous obesity pattern was therefore not reproduced by the secondary cardiometabolic indicators. Table 4 reports these estimates and their uncertainty.

**Table 4.**
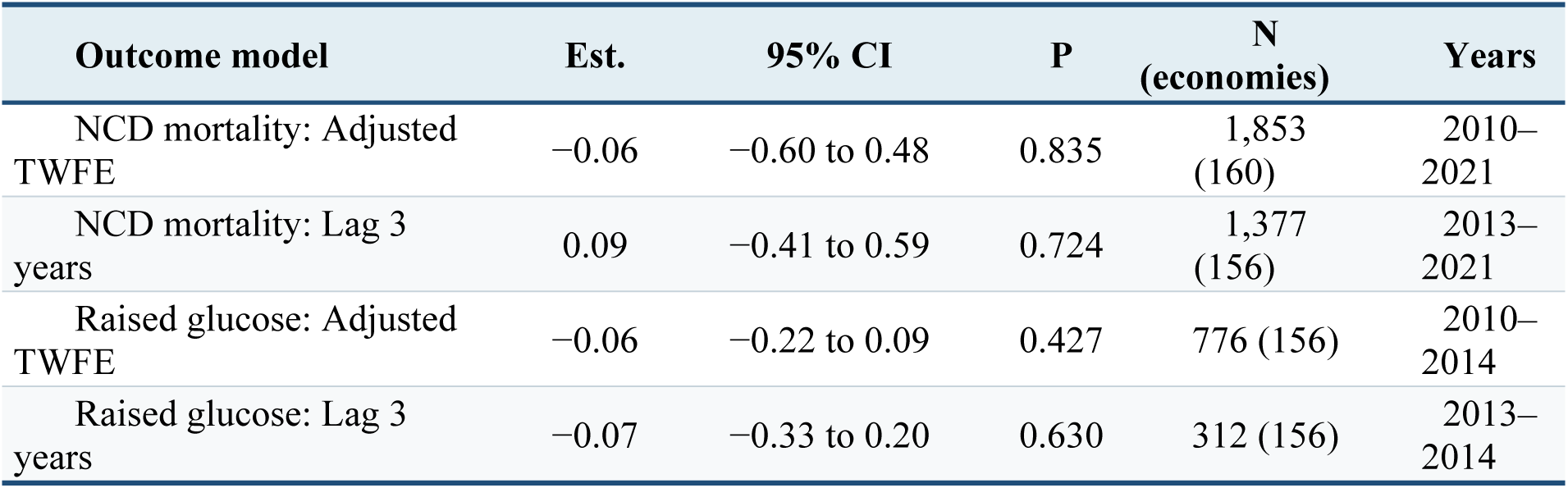
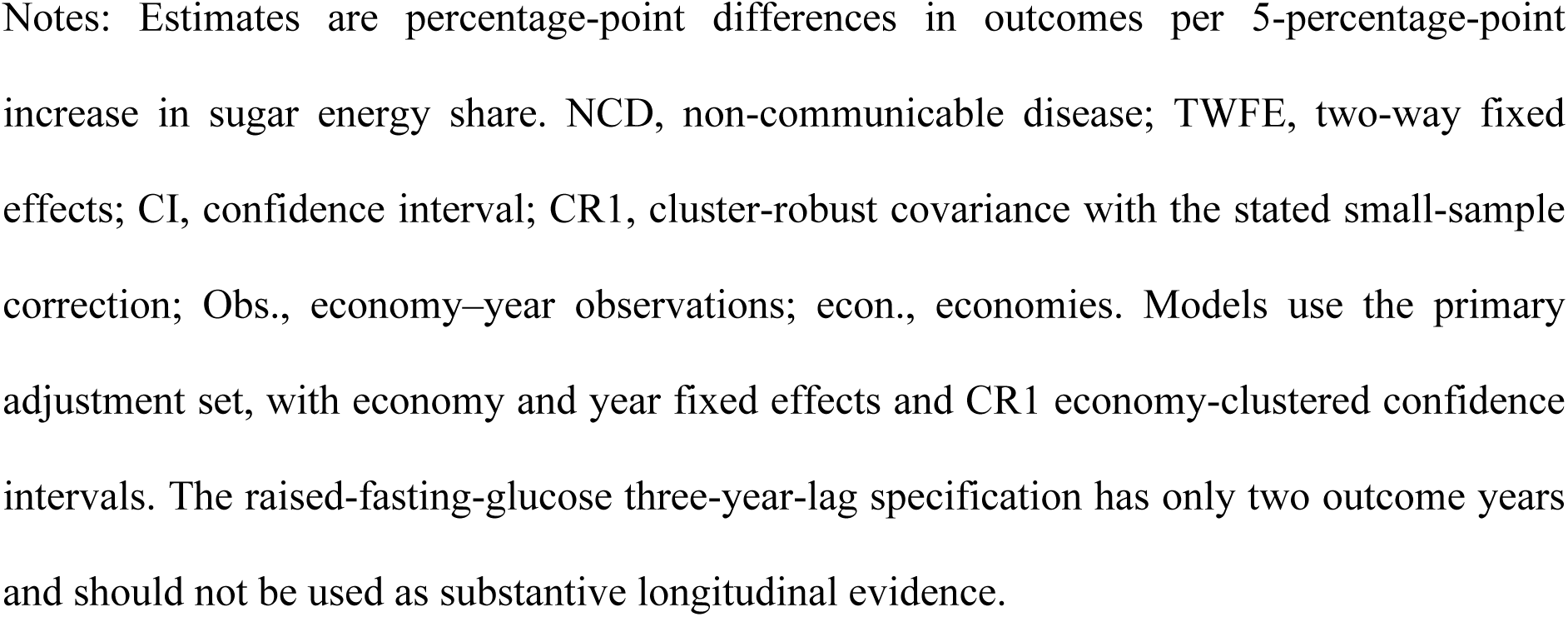
Adjusted associations with secondary cardiometabolic outcomes.

| Outcome model | Est. | 95% CI | P | N<br>(economies) | Years |
| --- | --- | --- | --- | --- | --- |
| NCD mortality: Adjusted TWFE | −0.06 | −0.60 to 0.48 | 0.835 | 1,853<br>(160) | 2010–2021 |
| NCD mortality: Lag 3 years | 0.09 | −0.41 to 0.59 | 0.724 | 1,377<br>(156) | 2013–2021 |
| Raised glucose: Adjusted TWFE | −0.06 | −0.22 to 0.09 | 0.427 | 776 (156) | 2010–2014 |
| Raised glucose: Lag 3 years | −0.07 | −0.33 to 0.20 | 0.630 | 312 (156) | 2013–2014 |

Notes: Estimates are percentage-point differences in outcomes per 5-percentage-point increase in sugar energy share. NCD, non-communicable disease; TWFE, two-way fixed effects; CI, confidence interval; CR1, cluster-robust covariance with the stated small-sample correction; Obs., economy–year observations; econ., economies. Models use the primary adjustment set, with economy and year fixed effects and CR1 economy-clustered confidence intervals. The raised-fasting-glucose three-year-lag specification has only two outcome years and should not be used as substantive longitudinal evidence.

## Discussion

### Principal findings

This global panel produced a coherent measurement diagnosis rather than evidence of a protective effect. Economies with higher long-run sugar and sweetener shares had higher WHO-modelled adult-obesity prevalence, whereas the adjusted contemporaneous within-economy estimate was negative. On a common set of observations, contemporaneous and lagged coefficients approached zero while the future-exposure association remained negative and differed formally from the contemporaneous and one-year-lag coefficients. The two-part ILR balance attenuated the inverse estimate to the boundary of conventional statistical significance. Economy-specific trends, population weighting, absolute exposure units and secondary outcomes also failed to reproduce a stable pattern. Independent re-estimation and wild-cluster-bootstrap inference verified the computation of the principal model, but computational validity did not rescue its temporal or construct interpretation. Collectively, the findings do not support a simple causal interpretation in either direction.

Earlier cross-national studies linked national sugar or soft-drink availability to diabetes and obesity, principally through comparisons among countries [13,14]. A recent Sub-Saharan African preprint similarly reported a positive between-country association that did not persist within countries [15]. In contrast, a Spanish study reported strong predictive performance from Food Balance Sheet nutrient supplies [16]. The present study extends this contested evidence globally and does not dispute previously reported between-economy patterns. It shows that those patterns answer a different question from whether changes in the same availability measure track changes in obesity within economies. Across the temporal, trend, weighting and exposure-unit diagnostics examined, the latter interpretation was not stable. The transferable contribution is that an ecological indicator must be evaluated against its precise estimand and decision use: cross-sectional agreement cannot license longitudinal interpretation.

The positive between-economy association is consistent with several explanations: greater availability of refined carbohydrates and processed foods, correlations with income and urbanisation, a broader dietary transition, differential measurement, or other features of national food systems. It is not an estimate of an individual effect. Likewise, the inverse within-economy coefficient does not imply that increasing sugar intake reduces obesity. Its disappearance for contemporaneous exposure in the common temporal sample, persistence for a future exposure, attenuation under the ILR balance, and dependence on trend and exposure unit are more consistent with trend structure, ratio behaviour, modelled outcomes, time-varying confounding and measurement error than with a protective effect.

### Why the sign reversal matters

Within–between sign reversal is scientifically informative because it shows that the data do not support a single interpretation across distinct economy-level comparisons. It is not, by itself, evidence that either coefficient is biased because the two models target different estimands. Cross-sectional levels largely reflect enduring differences among economies: the between SD of sugar share was nearly four times the within SD. The longitudinal estimate was therefore derived from relatively small deviations around persistent national levels. With limited and error-prone exposure variation, fixed-effects estimates can be attenuated or distorted, a documented problem in panel errors-in-variables settings [35]. Smooth outcome trajectories further make short-run contrasts sensitive to specification. The residual autoregressive coefficient of 0.93 indicates substantial persistence and limited independent within-economy information. Economy clustering addresses the covariance structure of inference but cannot establish temporal ordering or isolate the source of persistence.

The energy-share denominator adds another layer. A declining share can result from lower sugar kilocalories, higher energy availability from other commodities, or both. The formal two-part ILR balance modelled the relative information in sugar and the remainder of dietary energy without zeros or imputation [36]. Its median-equivalent five-point estimate remained negative but attenuated from −0.68 to −0.30, with a confidence interval crossing the null. This analysis is preferable to treating a bounded share as unconstrained. Still, it remains a coarse two-component composition because the masterfile does not partition non-sugar energy into substantively distinct food groups. When total energy is included as a covariate, both the original share and ILR estimates retain a substitution interpretation [37]. The near-zero mass estimate and imprecise absolute-kilocalorie estimate further show that the primary result was not scale-invariant. The post-2019 decline in the population-weighted share coincided with continuing increases in modelled obesity, but the study cannot separate pandemic-era supply changes, revisions, composition or other macro processes.

### Temporal diagnostics and causal restraint

A future exposure cannot cause an outcome measured three years earlier. Nevertheless, a persistent future value can correlate with earlier exposure and with omitted processes extending across time [24]. The common-sample analysis sharpened this diagnostic: the contemporaneous and lagged coefficients were near zero, while the future-exposure coefficient remained −0.63 and differed from the contemporaneous coefficient by −0.58 points. This is not a definitive negative control, but the contrast cannot be explained by different analytical rows. High correlations among the four exposure timings indicate persistence, yet persistence alone does not explain why the future coefficient was materially more negative on identical outcome years. Together with the near-zero economy-trend estimate, the matched result strengthens the inference that temporal structure or time-varying bias—not a stable protective relation—drives the original inverse coefficient.

This restraint is essential because the independent evidence base on sugar is stronger than that of this ecological proxy. WHO guidance, randomised evidence on dietary sugars and controlled work on dietary processing concern intake and interventions [2,3,5]. The present results neither overturn those findings nor evaluate a specific beverage tax, reformulation policy or individual dietary substitution. They show instead that national supply accounts are too remote from those constructs to adjudicate them alone.

### Implications for the socio-metabolic systems framework

The panel does not validate the behavioural, digital or recursive components of SMSF. It measured structural context, national food availability and modelled outcomes, but not perceived stress, executive control, household purchasing, time use, platform-level advertising exposure, biomarkers or metabolic mediation. Treating Internet penetration as algorithmic exposure, or long-hours prevalence as individual time scarcity, would substitute labels for measured constructs.

The inequality interaction is therefore hypothesis-generating only. Its direction could reflect the stage of the nutrition transition, differential under-recording, policy responses, denominator composition, sex-specific socioeconomic gradients or unmeasured food-system change; none is identified here. A direct SMSF test would require repeated dietary or purchase measures, validated stress and time-use indicators, platform-level exposure data, biomarkers and policy timing within a multilevel longitudinal design.

### Policy and surveillance implications

For surveillance, food-balance-sheet series remain valuable for describing food-system availability and long-run change [20]. Their labels should remain explicit: ‘availability’ or ‘supply’, not ‘consumption’. Estimates should be triangulated using household expenditure, retail scanner data, sales, waste adjustments and dietary surveys. For sugar and other national dietary-availability indicators, analysts should report both within- and between-economy estimands, use absolute and relative exposure units, probe leads and lags, and avoid ranking economies when input quality varies. A proxy that performs well for describing level differences should not be assumed to monitor change or evaluate policy without use-specific validation.

For policy, an unstable ecological proxy is not grounds for inaction. Fiscal, marketing, procurement and reformulation policies should be assessed using designs that exploit implementation timing and measure the intended purchase, sales or intake response. National supply aggregates can provide food-system context, but they should not be treated as policy exposures without policy-specific temporal data.

### Strengths and limitations

Strengths include a documented 217-economy frame; versioned official sources; explicit separation of source values, modelled estimates and derived proxies; no interpolation; exact calendar lags; within–between decomposition; economy and year fixed effects; economy-clustered and two-way-clustered uncertainty; maximal and common-sample temporal models with formal contrasts; economy-specific trends; difference models; population weighting; absolute-unit and formal ILR checks; secondary outcomes; multiple-testing correction for modifiers; independent estimator replication; wild-cluster-bootstrap inference; and an executable S1–S5 reproducibility package with checksums. The analysis treats contradictory results as evidence about identification rather than selecting the most favourable coefficient. Limitations fall into four domains. First, construct and measurement limitations constrain interpretation. The study is ecological and cannot recover individual exposure–outcome relations. FAOSTAT data include loss and waste and measure neither free-sugar intake nor sugar-sweetened-beverage consumption; annual national aggregation is also poorly aligned with individual dietary change and the long, heterogeneous latency of weight gain. WHO outcomes and SWIID values are modelled estimates, ILOSTAT working-time measures draw on heterogeneous national sources, Internet use is not digital marketing, and economy means suppress within-economy distributions. Provider uncertainty was not propagated, so the regression intervals are conditional and may understate total uncertainty. Second, temporal and statistical identification remain limited. Fixed effects remove time-invariant confounding but not time-varying diet, physical activity, education, policy, trade, conflict, price, reformulation or data-quality changes. Linear and additive models may average non-linear or heterogeneous relationships, while the three contextual interactions cover only a narrow subset of heterogeneity. Residual serial dependence was high, and 14 annual observations per economy provide limited leverage for gradual exposures. Economy-specific trends may absorb gradual effects, whereas difference models may amplify measurement error; both are diagnostic rather than definitive.

Third, selection and transportability are constrained by the unbalanced complete-case panel, which may select economies by statistical capacity, income or survey systems. Matching temporal specifications removes variation caused by changing row composition but restricts inference to 1,228 observations from 156 economies during 2013–2020; it cannot correct selection into that sample. The short raised-glucose series and small South Asia and North America samples further limit outcome- and region-specific inference. Fourth, the completed checks narrow but do not remove analytical uncertainty. The two-part ILR balance combines all non-sugar energy and cannot identify substitution among meaningful food groups. Independent computational replication and bootstrap inference verify estimation, not the absence of omitted variables, measurement error or functional-form misspecification. Multiple diagnostic models increase the opportunity for chance patterns, despite the structured audit and correction for the three exploratory modifier tests. Reproducibility depends on preserving the supplied S1–S5 files and executing S2 in the documented environment. The analysis remains associational and is not a substitute for individual trials or credible policy natural experiments.

## Conclusion

National sugar and sweetener availability showed a positive between-economy association with WHO-modelled adult-obesity prevalence. Still, it was not a temporally or construct-stable indicator of within-economy change across the economies and years examined. On identical observations, contemporaneous and lagged coefficients approached zero while the three-year-future coefficient remained negative and differed from the contemporaneous coefficient. Formal compositional treatment attenuated the inverse estimate and included the null; trend, weighting, absolute-unit and secondary-outcome checks also lacked coherence. Exact independent replication and wild-cluster-bootstrap inference confirmed that these contradictions were not coding artefacts. They must not be interpreted as evidence that sugar protects against obesity. Food-balance-sheet availability remains valuable for describing national food systems, but it cannot by itself validate individual intake effects, behavioural mechanisms or within-economy policy impacts. The principal contribution is a use-specific boundary of inference for ecological surveillance: between-economy ranking does not establish fitness for longitudinal tracking.

## Data availability

All data required to reproduce the reported findings are supplied as Supporting Information with this submission. S1 Data contains the harmonised economy–year workbook, codebook, coverage summary, provenance ledger, source records and an analysis quality-control sheet. S2 Code is the executable Python bundle. S3 Appendix supplies detailed methods and supplementary tables; S4 Checklist provides the completed STROBE mapping; and S5 Results contains machine-readable coefficients, contrasts, diagnostics, metadata snapshots, figure data, analysis summary and SHA-256 manifest. The original public data remain available from FAO; WHO GHO through the Our World in Data mirrors used for obesity and raised fasting glucose; WHO GHO via the World Bank and Our World in Data for premature NCD mortality; and the World Bank, SWIID and ILOSTAT at the cited locations. Provider terms and attribution requirements remain applicable.

## Code availability

The complete analysis code is provided as S2 Code and was executed successfully from the locked S1 workbook using Python 3.12.13 and the versions listed in requirements.txt. It reconstructs derived variables, fits every reported model, performs matched-sample Wald contrasts, estimates the two-part ILR balance, independently validates the principal fixed-effects and CR1 estimates, runs the 999-repetition wild-cluster bootstrap, regenerates figures and writes the S5 machine-readable outputs: the fixed random seed and SHA-256 manifest support deterministic verification.

## Author contributions

Z.A.N.: conceptualisation; methodology; data curation; formal analysis; software; validation; visualisation; writing—original draft; writing—review and editing; project administration.

## Data Availability

All data and code required to reproduce the analyses are supplied with this submission. S1 Data contains the harmonised economy-year dataset, data dictionary, coverage summary, provenance ledger, country crosswalk, merge quality-control records, filtered source records and analysis quality-control sheet. S2 Code contains the executable Python analysis code and locked package requirements. S3 Appendix provides the supplementary methods, diagnostics and tables. The original source data are publicly available from FAOSTAT, WHO Global Health Observatory series disseminated through Our World in Data, the World Bank World Development Indicators, the Standardised World Income Inequality Database and ILOSTAT through the links provided. No legal or ethical restrictions apply to the analytical data supplied with this submission.

https://ilostat.ilo.org/data/bulk/

https://doi.org/10.7910/DVN/LM4OWF

https://databank.worldbank.org/source/world-development-indicators

https://ourworldindata.org/grapher/diabetes-prevalence-who-gho

https://ourworldindata.org/grapher/mortality-from-ncds-sdgs

https://ourworldindata.org/grapher/obesity-prevalence-adults-who-gho

https://www.fao.org/faostat/en/

## Acknowledgements

The author acknowledges FAO, WHO, the World Bank, Our World in Data, Frederick Solt and the International Labour Organisation for maintaining and disseminating open international statistical resources.

## Supporting information captions

S1 Data. Harmonised analysis dataset. Locked economy–year workbook containing the master panel, data dictionary, coverage summary, provenance ledger, crosswalk, merge quality control, filtered source records and analysis quality-control sheet.

S2 Code. Executable reproducibility bundle. Python code and locked requirements used to derive variables, fit every reported model, conduct matched-sample contrasts, estimate the compositional balance, validate the estimator, run the wild-cluster bootstrap, export diagnostics and regenerate figures.

S3 Appendix. Supplementary methods and tables. Detailed integration and estimation procedures, sample cascade, complete matched temporal coefficients and Wald contrasts, compositional analysis, independent validation, bootstrap sensitivity and additional diagnostics.

S4 Checklist. STROBE reporting checklist—completed reporting-location checklist adapted to this ecological longitudinal observational study.

S5 Results. Machine-readable analytical outputs. Full-precision coefficients, matched contrasts, compositional results, validation and bootstrap statistics, diagnostic files, metadata snapshots, figure data, analysis summary and SHA-256 manifest.

