## Supplementary material for "Inferential instability of national sugar and sweetener availability as an indicator of adult obesity trajectories: A global within–between panel audit": S3 Appendix

### S3 Appendix. Supplementary methods and tables

This appendix documents the completed analyses added before submission. It should be read alongside S1 Data, S2 Code and S5 Results. All reported values were generated by running S2 Code against the locked S1 workbook; the CSV files in S5 preserve full numerical precision.

#### Supplementary Methods S1. Data integration and quality control

The unit of analysis was the economy or territory by calendar year. The locked frame contained 3,038 rows for 217 economies and territories from 2010 to 2023. Source records from FAOSTAT, the World Health Organization, the World Bank, SWIID and ILOSTAT were joined using the ISO 3166-1 alpha-3 code and the exact calendar year. Aggregate regions and income groups were excluded. Duplicate ISO-year keys were prohibited. No interpolation, carry-forward or model-based imputation was used. S1 Data contains the master panel, coverage and codebook sheets, provenance ledger, country crosswalk, merge quality control and filtered source records. The added Analysis_QC sheet records the executed analytical samples and package checks without altering source values.

#### Supplementary Methods S2. Principal fixed-effects estimator

The principal model regressed adult obesity prevalence on sugar energy share, scaled by five percentage points, and on the prespecified time-varying controls, economy fixed effects and year fixed effects. The controls were total dietary energy supply, log GDP per capita, urban population share, unemployment, Internet use, population aged 65 years or older, and current health expenditure as a share of GDP. Ordinary least squares was estimated using the full design matrix. The covariance used an economy-clustered CR1 sandwich correction, with inference based on $G - 1$ cluster degrees of freedom. A two-way economy-and-year-clustered sensitivity model and the prespecified trend, difference, weighting, exposure-unit and sample diagnostics were reproduced from the same locked data.

#### Supplementary Methods S3. Common-sample temporal analysis

The common temporal sample required adult obesity at outcome year t; sugar energy share at t, $t - 1$, $t - 3$ and $t + 3$; and every contemporaneous covariate to be observed simultaneously. This produced 1,228 outcome-year observations from 156 economies during 2013–2020. Four economy-and-year fixed-effects models were fitted to exactly these rows, with only the temporal exposure column varying. To compare coefficients formally, economy-level score vectors from all four equations were stacked. A block-diagonal matrix of model-specific bread matrices premultiplied and postmultiplied the stacked cluster meat; the common CR1 factor was then applied. Pairwise Wald contrasts used this cross-equation covariance and 155 cluster degrees of freedom. Thus, differences in coefficients cannot be attributed to changes in the set of economies, outcome years, or covariate observations.

**Supplementary Methods S4. Compositional sensitivity analysis**

The dietary energy composition was partitioned into sugar-and-sweetener kilocalories and all remaining dietary energy kilocalories. The isometric log-ratio balance was $z = \sqrt{(1/2)} \times \ln[\text{sugar kilocalories}/(\text{total kilocalories} - \text{sugar kilocalories})]$. Both components were strictly positive in every primary analytical row; consequently, no pseudocount, multiplicative replacement, or other zero-handling was applied. The fixed-effects model replaced the bounded sugar share with z and retained total energy and all other controls. The primary compositional estimand was the outcome difference per one-unit increase in z. For comparability with the five-percentage-point share contrast, the coefficient was also multiplied by $\Delta z = 0.332088$, the balance change from the analytical median share of 9.789% to 14.789%. This two-part balance is formally coherent for sugar versus the remainder, although it cannot distinguish substitutions among food groups within the remainder.

**Supplementary Methods S5. Independent validation and bootstrap inference**

The full-dummy fixed-effects coefficient was independently re-estimated with scikit-learn LinearRegression using no additional intercept. Residuals and economy score vectors were then recomputed independently, and the CR1 covariance was reconstructed from those scores. Agreement with the matrix implementation was required within $1 \times 10^{-10}$ for both the sugar coefficient and its standard error. A restricted Rademacher wild-cluster-bootstrap-t test evaluated the null of no principal sugar association using 999 repetitions and seed 20260811. The bootstrap used economy-level sign weights, imposed the null in the residualised equation and studentised each replicate with its clustered standard error.

**Supplementary Methods S6. Reproducible execution**

The verified environment used Python 3.12.13, pandas 2.2.3, NumPy 2.3.5, SciPy 1.17.0, scikit-learn 1.8.0, Matplotlib 3.10.8, Pillow 12.2.0 and openpyxl 3.1.5. S2 Code includes a locked requirements file and one command-line entry point. Each clean run writes model coefficients, matched contrasts, correlations, compositional estimates, validation and bootstrap statistics, descriptive summaries, metadata snapshots, figure data, TIFF figures, analysis_summary.json and analysis_manifest.csv. The manifest records file sizes and SHA-256 checksums.

**Table S1. Analytical sample cascade**

| Step | Observations | Economies | Years | % of full frame |
|---|---|---|---|---|
| Full economy-year frame | 3,038 | 217 | 2010–2023 | 100.0% |
| Sugar energy share observed | 2,397 | 177 | 2010–2023 | 78.9% |
| Sugar share and adult obesity observed | 2,355 | 174 | 2010–2023 | 77.5% |
| Primary adjusted complete cases | 2,153 | 160 | 2010–2023 | 70.9% |
| Matched temporal complete cases | 1,228 | 156 | 2013–2020 | 40.4% |
| Compositional complete cases | 2,153 | 160 | 2010–2023 | 70.9% |
| Inequality interaction complete cases | 1,727 | 150 | 2010–2023 | 56.8% |
| Working-time interaction complete cases | 1,248 | 142 | 2010–2023 | 41.1% |
| NCD mortality adjusted complete cases | 1,853 | 160 | 2010–2021 | 61.0% |
| Raised-glucose adjusted complete cases | 776 | 156 | 2010–2014 | 25.5% |

Notes: Samples are model-specific and are not necessarily nested. The matched temporal sample requires all four exposure timings and every primary covariate simultaneously.

**Table S2A. Common-sample temporal coefficients**

| Temporal exposure | Estimate | SE | 95% CI | P | Obs. | Econ. | Years |
|---|---|---|---|---|---|---|---|
| contemporaneous | −0.06 | 0.184 | −0.42 to 0.31 | 0.760 | 1,228 | 156 | 2013–2020 |
| lag 1 year | −0.08 | 0.177 | −0.42 to 0.27 | 0.671 | 1,228 | 156 | 2013–2020 |
| lag 3 years | −0.05 | 0.209 | −0.46 to 0.36 | 0.809 | 1,228 | 156 | 2013–2020 |
| Future exposure (lead 3) | −0.63 | 0.191 | −1.01 to −0.26 | 0.001 | 1,228 | 156 | 2013–2020 |

Notes: Outcome is adult obesity prevalence. Every estimate is per a five-percentage-point higher sugar energy share and uses the same observations and controls.

**Table S2B. Cluster-aware Wald contrasts among common-sample coefficients**

| Contrast | Difference | SE | 95% CI | P |
|---|---|---|---|---|
| lag 1 year minus contemporaneous | −0.02 | 0.121 | −0.26 to 0.22 | 0.876 |
| lag 3 years minus contemporaneous | 0.01 | 0.249 | −0.49 to 0.50 | 0.982 |
| future exposure (lead 3) minus contemporaneous | −0.58 | 0.186 | −0.95 to −0.21 | 0.002 |
| lag 3 years minus lag 1 year | 0.02 | 0.175 | −0.32 to 0.37 | 0.888 |
| future exposure (lead 3) minus lag 1 year | −0.56 | 0.247 | −1.05 to −0.07 | 0.025 |
| future exposure (lead 3) minus lag 3 years | −0.58 | 0.309 | −1.19 to 0.03 | 0.060 |

Notes: Differences are second-listed minus first-listed coefficients. The stacked economy-clustered covariance accounts for the fact that all equations use the same rows.

The contemporaneous and lagged coefficients were near zero on the common sample. The future-exposure coefficient remained negative and differed from the contemporaneous coefficient (P = 0.002) and one-year-lag coefficient (P = 0.025). Matching, therefore, strengthened the conclusion of temporal instability: the original inverse contemporaneous result disappeared, whereas an exposure measured three years after the outcome retained a materially negative association.

**Table S3. Formal two-part compositional sensitivity analysis**

| Estimand | Scaling | Estimate | SE | 95% CI | P | Obs. (econ.) |
|---|---|---|---|---|---|---|
| sugar-to-remainder balance | per one-unit increase in sqrt(1/2) × ln(sugar kcal/non-sugar kcal) | −0.909 | 0.462 | −1.821 to 0.003 | 0.051 | 2,153 (160) |
| balance: median-equivalent 5-point contrast | ILR change 0.332088, equivalent to 9.789% to 14.789% | −0.302 | 0.153 | −0.605 to 0.001 | 0.051 | 2,153 (160) |

Notes: ILR, isometric log-ratio. Zero replacement was not used because sugar and non-sugar energy were strictly positive in all 2,153 analytical observations.

The compositional coefficient retained an inverse direction, but the median-equivalent magnitude attenuated to −0.302 percentage points, and its interval included the null. Thus, the conventional $P < 0.05$ inference from the bounded energy-share model did not survive formal log-ratio treatment, while the broader conclusion of construct instability did.

**Table S4. Independent estimator validation and wild-cluster-bootstrap sensitivity**

| Check | Estimate | SE | Interval/quantiles | P | Verification |
|---|---|---|---|---|---|
| Independent fixed-effects replication | −0.684 | 0.252 | −1.181 to −0.186 | 0.007 | Coefficient Δ = 0; SE Δ = $5.25 \times 10^{-12}$; both tolerances passed |
| Restricted wild-cluster-bootstrap-t | −0.684 | 0.252 | Bootstrap t quantiles: −1.974 to 2.031 | 0.010 | 999 repetitions; seed 20260811; 160 clusters |

Notes: The independent implementation used scikit-learn full-dummy fixed effects and an independently aggregated CR1 covariance. The bootstrap P value is the finite-repetition two-sided value with the plus-one correction.

**Table S5. Reproduced adult-obesity estimates and diagnostics**

| Model / estimand | Estimate | 95% CI | P | Obs. | Econ. | Years |
|---|---|---|---|---|---|---|
| Between-economy component | 2.373 | 0.516 to 4.230 | 0.013 | 2,153 | 160 | 2010–2023 |
| Within-economy component | −0.578 | −1.605 to 0.450 | 0.269 | 2,153 | 160 | 2010–2023 |
| Exposure-only TWFE | −0.643 | −1.147 to −0.138 | 0.013 | 2,355 | 174 | 2010–2023 |
| Adjusted TWFE | −0.684 | −1.181 to −0.186 | 0.007 | 2,153 | 160 | 2010–2023 |
| Lag 1 year | −0.541 | −1.015 to −0.067 | 0.026 | 1,995 | 160 | 2011–2023 |
| Lag 3 years | −0.206 | −0.636 to 0.224 | 0.345 | 1,677 | 160 | 2013–2023 |
| Future exposure (lead 3) | −0.671 | −1.081 to −0.262 | 0.001 | 1,700 | 160 | 2010–2020 |
| Economy-specific trends | −0.026 | −0.105 to 0.053 | 0.519 | 2,153 | 160 | 2010–2023 |
| Annual difference | −0.082 | −0.136 to −0.029 | 0.003 | 1,990 | 160 | 2011–2023 |
| Five-year difference | −0.455 | −0.838 to −0.072 | 0.020 | 1,351 | 155 | 2015–2023 |
| Population-weighted TWFE | −0.351 | −1.155 to 0.453 | 0.390 | 2,153 | 160 | 2010–2023 |
| Two-way clustered SE | −0.684 | −1.350 to −0.017 | 0.045 | 2,153 | 160 | 2010–2023 |
| At least 10 complete years | −0.697 | −1.196 to −0.197 | 0.007 | 2,106 | 152 | 2010–2023 |
| Microstate/outlier exclusion | −0.553 | −1.084 to −0.022 | 0.041 | 1,898 | 139 | 2010–2023 |

| Model / estimand | Estimate | 95% CI | P | Obs. | Econ. | Years |
|---|---|---|---|---|---|---|
| Pre-2020 period | −0.203 | −0.606 to 0.200 | 0.321 | 1,545 | 160 | 2010–2019 |
| Sugar supply mass (per 10 kg) | −0.023 | −0.212 to 0.165 | 0.808 | 2,153 | 160 | 2010–2023 |
| Sugar energy (per 100 kcal) | −0.338 | −0.700 to 0.023 | 0.066 | 2,153 | 160 | 2010–2023 |

Notes: Scaling is per 5-percentage-point increase in sugar energy share, except where the model label specifies per 10 kg/person/year or per 100 kcal/person/day. All values match the manuscript after rounding.

**Table S6. Structural-context interactions**

| Context | Mean (between SD) | Interaction | 95% CI | P | q | Obs. (econ.) |
|---|---|---|---|---|---|---|
| Income inequality | 38.58 (7.76) | −0.659 | −1.115 to −0.203 | 0.005 | 0.015 | 1,727 (150) |
| Long-hours work | 19.31 (12.27) | −0.108 | −0.741 to 0.525 | 0.737 | 0.737 | 1,248 (142) |
| Digital access | 52.47 (27.10) | 0.155 | −0.317 to 0.627 | 0.518 | 0.737 | 2,153 (160) |

Notes: q values use the Benjamini–Hochberg procedure across the three prespecified interaction tests.

**Table S7. Secondary-outcome estimates**

| Outcome model | Estimate | 95% CI | P | Obs. (econ.) | Years |
|---|---|---|---|---|---|
| NCD mortality: adjusted TWFE | −0.057 | −0.595 to 0.481 | 0.835 | 1,853 (160) | 2010–2021 |
| NCD mortality: lag 3 years | 0.089 | −0.410 to 0.589 | 0.724 | 1,377 (156) | 2013–2021 |
| Raised glucose: adjusted TWFE | −0.063 | −0.219 to 0.093 | 0.427 | 776 (156) | 2010–2014 |
| Raised glucose: lag 3 years | −0.065 | −0.334 to 0.203 | 0.630 | 312 (156) | 2013–2014 |

Notes: The raised-fasting-glucose three-year-lag specification contains outcome years 2013–2014 only and is retained for completeness.

**Table S8. Correlations among temporal exposures in the common sample**

| Exposure | Current | Lag 1 | Lag 3 | Lead 3 |
|---|---|---|---|---|
| Current | 1.000 | 0.978 | 0.961 | 0.949 |
| Lag 1 | 0.978 | 1.000 | 0.974 | 0.939 |
| Lag 3 | 0.961 | 0.974 | 1.000 | 0.927 |
| Lead 3 | 0.949 | 0.939 | 0.927 | 1.000 |

Notes: Pearson correlations are calculated across the 1,228 common-sample outcome-year observations. High persistence motivates caution but does not invalidate the cross-equation contrasts.

**Supplementary conclusion**

The completed checks resolve the previously identified analytical and reproducibility gaps. The matched analysis rules out changing row composition as the sole explanation for temporal inconsistency; the compositional model shows that the primary ratio-based inference is not robust at the conventional threshold; and independent replication plus wild-cluster bootstrap verifies the computation of the principal estimate. These results strengthen, rather than soften, the manuscript's central conclusion that national sugar availability is not a stable within-economy indicator of adult-obesity trajectories.
