## Supplementary material for "Inferential instability of national sugar and sweetener availability as an indicator of adult obesity trajectories: A global within–between panel audit": S2 Code: analysis.py

```
#!/usr/bin/env python3
"""Reproduce and extend the national sugar-availability panel audit.

The script reads S1_Data.xlsx, recreates the reported estimates, conducts the
matched-sample temporal and two-part compositional analyses, independently
validates the fixed-effects fit with scikit-learn, and performs a restricted
wild-cluster-bootstrap-t sensitivity analysis.
"""

from __future__ import annotations

import argparse
import hashlib
import json
import math
import os
from dataclasses import dataclass
from pathlib import Path

os.environ.setdefault("MPLCONFIGDIR", "/tmp/sugar_ncd_mplconfig")

import matplotlib
matplotlib.use("Agg")
import matplotlib.pyplot as plt
from matplotlib.patches import FancyArrowPatch, FancyBboxPatch
import numpy as np
import pandas as pd
import scipy
from scipy import linalg, stats
from sklearn.linear_model import LinearRegression
from PIL import Image


# PLOS figure typography: Arial-family text throughout. Nimbus Sans is the
# metrically compatible fallback in the verified execution environment.
plt.rcParams.update(
    {
        "font.family": "sans-serif",
        "font.sans-serif": ["Arial", "Nimbus Sans", "Liberation Sans"],
        "mathtext.fontset": "dejavusans",
        "axes.unicode_minus": True,
        "axes.linewidth": 0.8,
        "xtick.major.width": 0.8,
        "ytick.major.width": 0.8,
    }
)


CONTROL_VARS = [
    "energy100",
    "log_gdp",
    "urban10",
    "unemployment5",
    "internet10",
    "age65_5",
    "health_expenditure_gdp_pct",
]


def benjamini_hochberg(p_values: pd.Series) -> np.ndarray:
    """Return monotone Benjamini-Hochberg adjusted P values."""
    p = np.asarray(p_values, dtype=float)
    order = np.argsort(p)
    ranked = p[order]
    adjusted = ranked * len(p) / np.arange(1, len(p) + 1)
    adjusted = np.minimum.accumulate(adjusted[::-1])[::-1]
    out = np.empty_like(adjusted)
    out[order] = np.clip(adjusted, 0.0, 1.0)
    return out


def export_documentation_sheets(data_path: Path, output_path: Path) -> None:
    """Export the principal metadata sheets as portable CSV files."""
    for sheet in ["Coverage", "Codebook", "Provenance", "Country_Crosswalk", "Merge_QC"]:
        frame = pd.read_excel(data_path, sheet_name=sheet, header=3)
        frame = frame.dropna(how="all").dropna(axis=1, how="all")
        frame.to_csv(output_path / f"metadata_{sheet.lower()}.csv", index=False)


def write_manifest(output_path: Path) -> None:
    """Record byte sizes and SHA-256 checksums for generated analytical outputs."""
    rows = []
    for path in sorted(output_path.iterdir()):
        if path.is_file() and path.name != "analysis_manifest.csv":
            digest = hashlib.sha256(path.read_bytes()).hexdigest()
            rows.append({"file": path.name, "bytes": path.stat().st_size, "sha256": digest})
    pd.DataFrame(rows).to_csv(output_path / "analysis_manifest.csv", index=False)


@dataclass
class Fit:
    label: str
    data: pd.DataFrame
    y: np.ndarray
    X: np.ndarray
    names: list[str]
    beta: np.ndarray
    residual: np.ndarray
    covariance: np.ndarray
    standard_error: np.ndarray
    nobs: int
    rank: int
    groups: int
    df_resid: int
    cr1_factor: float
    weights: np.ndarray | None = None

    def result(self, term: str, family: str = "", scale_note: str = "") -> dict:
        j = self.names.index(term)
        estimate = float(self.beta[j])
        se = float(self.standard_error[j])
        z = estimate / se
        cluster_df = self.groups - 1
        p = float(2 * stats.t.sf(abs(z), cluster_df))
        critical = float(stats.t.ppf(0.975, cluster_df))
        lo, hi = estimate - critical * se, estimate + critical * se
        return {
            "family": family,
            "model": self.label,
            "term": term,
            "scale": scale_note,
            "estimate": estimate,
            "standard_error": se,
            "ci_lower": float(lo),
            "ci_upper": float(hi),
            "p_value": p,
            "observations": self.nobs,
            "economies": self.groups,
            "cluster_df": cluster_df,
            "first_year": int(self.data["year"].min()),
            "last_year": int(self.data["year"].max()),
            "rank": self.rank,
            "df_residual": self.df_resid,
            "cr1_factor": self.cr1_factor,
        }


def read_masterfile(path: Path) -> pd.DataFrame:
    df = pd.read_excel(path, sheet_name="Master_CountryYear", header=3)
    df = df[df["iso3"].notna() & df["year"].notna()].copy()
    df["year"] = pd.to_numeric(df["year"], errors="raise").astype(int)
    numeric = [c for c in df.columns if c not in {"iso3", "country_name", "region", "income_group"}]
    for col in numeric:
        df[col] = pd.to_numeric(df[col], errors="coerce")
    if df.duplicated(["iso3", "year"]).any():
        raise ValueError("Duplicate iso3-year keys detected")

    df["sugar5"] = df["sugar_energy_share_pct"] / 5.0
    df["sugar_kg10"] = df["sugar_supply_kg_capita_year"] / 10.0
    df["sugar_kcal100"] = df["sugar_supply_kcal_capita_day"] / 100.0
    df["energy100"] = df["total_dietary_energy_kcal_capita_day"] / 100.0
    df["log_gdp"] = np.log(df["gdp_per_capita_ppp_const_2021_intl"].where(lambda x: x > 0))
    df["urban10"] = df["urban_population_pct"] / 10.0
    df["unemployment5"] = df["unemployment_pct"] / 5.0
    df["internet10"] = df["internet_users_pct"] / 10.0
    df["age65_5"] = df["population_age65_plus_pct"] / 5.0

    nonsugar = df["total_dietary_energy_kcal_capita_day"] - df["sugar_supply_kcal_capita_day"]
    valid = (df["sugar_supply_kcal_capita_day"] > 0) & (nonsugar > 0)
    df["ilr_sugar_remainder"] = np.nan
    df.loc[valid, "ilr_sugar_remainder"] = (
        math.sqrt(0.5)
        * np.log(df.loc[valid, "sugar_supply_kcal_capita_day"] / nonsugar.loc[valid])
    )

    source = df[["iso3", "year", "sugar5"]].copy()
    for name, offset in [("sugar5_lag1", 1), ("sugar5_lag3", 3), ("sugar5_lead3", -3)]:
        shifted = source.rename(columns={"sugar5": name}).copy()
        shifted["year"] = shifted["year"] + offset
        df = df.merge(shifted, on=["iso3", "year"], how="left", validate="one_to_one")
    return df


def make_design(
    data: pd.DataFrame,
    xvars: list[str],
    *,
    economy_fe: bool = True,
    year_fe: bool = True,
    region_fe: bool = False,
    economy_trends: bool = False,
) -> tuple[np.ndarray, list[str]]:
    blocks = [np.ones((len(data), 1)), data[xvars].to_numpy(float)]
    names = ["Intercept", *xvars]
    country_dummies = None
    if economy_fe:
        country_dummies = pd.get_dummies(data["iso3"], prefix="economy", drop_first=True, dtype=float)
        blocks.append(country_dummies.to_numpy())
        names.extend(country_dummies.columns.tolist())
    if year_fe:
        year_dummies = pd.get_dummies(data["year"], prefix="year", drop_first=True, dtype=float)
        blocks.append(year_dummies.to_numpy())
        names.extend(year_dummies.columns.tolist())
    if region_fe:
        region_dummies = pd.get_dummies(data["region"], prefix="region", drop_first=True, dtype=float)
        blocks.append(region_dummies.to_numpy())
        names.extend(region_dummies.columns.tolist())
    if economy_trends:
        centred_year = (data["year"] - data["year"].mean()).to_numpy(float)
        all_countries = pd.get_dummies(data["iso3"], prefix="trend", drop_first=False, dtype=float)
        trend_block = all_countries.to_numpy() * centred_year[:, None]
        blocks.append(trend_block)
        names.extend(all_countries.columns.tolist())
    return np.column_stack(blocks), names


def cluster_meat(X: np.ndarray, residual: np.ndarray, groups: np.ndarray) -> np.ndarray:
    meat = np.zeros((X.shape[1], X.shape[1]), dtype=float)
    for group in pd.unique(groups):
        mask = groups == group
        score = X[mask].T @ residual[mask]
        meat += np.outer(score, score)
    return meat


def fit_clustered(
    data: pd.DataFrame,
    yvar: str,
    xvars: list[str],
    label: str,
    *,
    economy_fe: bool = True,
    year_fe: bool = True,
    region_fe: bool = False,
    economy_trends: bool = False,
    weights: str | None = None,
) -> Fit:
    required = [yvar, *xvars, "iso3", "year"]
    if region_fe:
        required.append("region")
    if weights:
        required.append(weights)
    d = data.dropna(subset=required).copy().reset_index(drop=True)
    X, names = make_design(
        d,
        xvars,
        economy_fe=economy_fe,
        year_fe=year_fe,
        region_fe=region_fe,
        economy_trends=economy_trends,
    )
    y = d[yvar].to_numpy(float)
    w = None
    if weights:
        w = d[weights].to_numpy(float)
        w = w / np.nanmean(w)
        root_w = np.sqrt(w)
        X_fit = X * root_w[:, None]
        y_fit = y * root_w
    else:
        X_fit, y_fit = X, y

    beta, _, rank, _ = np.linalg.lstsq(X_fit, y_fit, rcond=None)
    residual_original = y - X @ beta
    residual_fit = y_fit - X_fit @ beta
    bread = np.linalg.pinv(X_fit.T @ X_fit, hermitian=True)
    groups = d["iso3"].to_numpy()
    meat = cluster_meat(X_fit, residual_fit, groups)
    nobs, groups_n = len(d), d["iso3"].nunique()
    df_resid = nobs - int(rank)
    cr1 = (groups_n / (groups_n - 1)) * ((nobs - 1) / df_resid)
    covariance = cr1 * bread @ meat @ bread
    se = np.sqrt(np.clip(np.diag(covariance), 0, None))
    return Fit(
        label=label,
        data=d,
        y=y,
        X=X,
        names=names,
        beta=beta,
        residual=residual_original,
        covariance=covariance,
        standard_error=se,
        nobs=nobs,
        rank=int(rank),
        groups=groups_n,
        df_resid=df_resid,
        cr1_factor=float(cr1),
        weights=w,
    )


def fit_explicit_design(data: pd.DataFrame, yvar: str, X: np.ndarray, names: list[str], label: str) -> Fit:
    d = data.reset_index(drop=True).copy()
    y = d[yvar].to_numpy(float)
    beta, _, rank, _ = np.linalg.lstsq(X, y, rcond=None)
    residual = y - X @ beta
    bread = np.linalg.pinv(X.T @ X, hermitian=True)
    groups = d["iso3"].to_numpy()
    meat = cluster_meat(X, residual, groups)
    nobs, groups_n = len(d), d["iso3"].nunique()
    df_resid = nobs - int(rank)
    cr1 = (groups_n / (groups_n - 1)) * ((nobs - 1) / df_resid)
    covariance = cr1 * bread @ meat @ bread
    se = np.sqrt(np.clip(np.diag(covariance), 0, None))
    return Fit(label, d, y, X, names, beta, residual, covariance, se, nobs, int(rank), groups_n, df_resid, float(cr1))


def mundlak_model(df: pd.DataFrame) -> tuple[Fit, list[dict]]:
    variables = ["sugar5", *CONTROL_VARS]
    # Period means are defined from every available observation of each
    # variable before complete-case restriction, as in the reported analysis.
    for var in variables:
        df[f"{var}_mean_full"] = df.groupby("iso3")[var].transform("mean")
        df[f"{var}_within_full"] = df[var] - df[f"{var}_mean_full"]
    required = ["obesity_adults_pct", *variables, "iso3", "year", "region"]
    d = df.dropna(subset=required).copy().reset_index(drop=True)
    xvars = []
    for var in variables:
        mean_name, within_name = f"{var}_mean_full", f"{var}_within_full"
        xvars.extend([mean_name, within_name])
    X, names = make_design(d, xvars, economy_fe=False, year_fe=True, region_fe=True)
    fit = fit_explicit_design(d, "obesity_adults_pct", X, names, "Within-between decomposition")
    rows = [
        fit.result("sugar5_mean_full", "within_between", "per 5-percentage-point higher economy mean"),
        fit.result("sugar5_within_full", "within_between", "per 5-percentage-point within-economy deviation"),
    ]
    rows[0]["model"] = "Between-economy component"
    rows[1]["model"] = "Within-economy component"
    return fit, rows


def matched_temporal(df: pd.DataFrame) -> tuple[list[Fit], pd.DataFrame, pd.DataFrame]:
    exposures = ["sugar5", "sugar5_lag1", "sugar5_lag3", "sugar5_lead3"]
    labels = ["Matched contemporaneous", "Matched lag 1 year", "Matched lag 3 years", "Matched future exposure (lead 3)"]
    required = ["obesity_adults_pct", *exposures, *CONTROL_VARS, "iso3", "year"]
    matched = df.dropna(subset=required).copy().reset_index(drop=True)
    fits = []
    rows = []
    for exposure, label in zip(exposures, labels):
        fit = fit_clustered(matched, "obesity_adults_pct", [exposure, *CONTROL_VARS], label)
        fits.append(fit)
        rows.append(fit.result(exposure, "matched_temporal", "per 5-percentage-point higher sugar energy share"))

    # Cluster-aware cross-equation covariance from stacked cluster score vectors.
    p = fits[0].X.shape[1]
    if any(f.X.shape[1] != p for f in fits):
        raise RuntimeError("Matched models do not share a common parameter dimension")
    bread = [np.linalg.pinv(f.X.T @ f.X, hermitian=True) for f in fits]
    groups = matched["iso3"].to_numpy()
    stacked_meat = np.zeros((p * len(fits), p * len(fits)))
    for group in pd.unique(groups):
        mask = groups == group
        scores = [f.X[mask].T @ f.residual[mask] for f in fits]
        stacked = np.concatenate(scores)
        stacked_meat += np.outer(stacked, stacked)
    block_bread = linalg.block_diag(*bread)
    cr1 = fits[0].cr1_factor
    stacked_cov = cr1 * block_bread @ stacked_meat @ block_bread
    indices = [m * p + fits[m].names.index(exposures[m]) for m in range(len(fits))]
    contrasts = []
    for i in range(len(fits)):
        for j in range(i + 1, len(fits)):
            a, b = indices[j], indices[i]
            diff = float(fits[j].beta[fits[j].names.index(exposures[j])] - fits[i].beta[fits[i].names.index(exposures[i])])
            variance = float(stacked_cov[a, a] + stacked_cov[b, b] - 2 * stacked_cov[a, b])
            se = math.sqrt(max(variance, 0))
            statistic = diff / se
            df_cluster = fits[0].groups - 1
            p_value = float(2 * stats.t.sf(abs(statistic), df_cluster))
            crit = float(stats.t.ppf(0.975, df_cluster))
            contrasts.append({
                "contrast": f"{labels[j]} minus {labels[i]}",
                "difference": diff,
                "standard_error": se,
                "ci_lower": diff - crit * se,
                "ci_upper": diff + crit * se,
                "t_statistic": statistic,
                "cluster_df": df_cluster,
                "p_value": p_value,
                "observations": len(matched),
                "economies": matched["iso3"].nunique(),
                "first_year": int(matched["year"].min()),
                "last_year": int(matched["year"].max()),
            })
    return fits, pd.DataFrame(rows), pd.DataFrame(contrasts)


def compositional_model(df: pd.DataFrame) -> tuple[Fit, pd.DataFrame, dict]:
    fit = fit_clustered(
        df,
        "obesity_adults_pct",
        ["ilr_sugar_remainder", *CONTROL_VARS],
        "Two-part ILR sugar-to-remainder balance",
    )
    base = fit.result(
        "ilr_sugar_remainder",
        "compositional",
        "per one-unit increase in sqrt(1/2) × ln(sugar kcal/non-sugar kcal)",
    )
    median_share = float(fit.data["sugar_energy_share_pct"].median())
    if median_share + 5 >= 100:
        raise ValueError("Invalid five-point compositional contrast")
    ilr_at_median = math.sqrt(0.5) * math.log(median_share / (100 - median_share))
    ilr_at_plus5 = math.sqrt(0.5) * math.log((median_share + 5) / (100 - median_share - 5))
    delta = ilr_at_plus5 - ilr_at_median
    scaled = base.copy()
    scaled.update({
        "model": "Two-part ILR balance: median-equivalent 5-point contrast",
        "scale": f"ILR change {delta:.6f}, equivalent to {median_share:.3f}% to {median_share + 5:.3f}%",
        "estimate": base["estimate"] * delta,
        "standard_error": base["standard_error"] * delta,
        "ci_lower": base["ci_lower"] * delta,
        "ci_upper": base["ci_upper"] * delta,
    })
    rows = pd.DataFrame([base, scaled])
    qc = {
        "analytic_observations": fit.nobs,
        "economies": fit.groups,
        "nonpositive_sugar_values": int((fit.data["sugar_supply_kcal_capita_day"] <= 0).sum()),
        "nonpositive_remainder_values": int(((fit.data["total_dietary_energy_kcal_capita_day"] - fit.data["sugar_supply_kcal_capita_day"]) <= 0).sum()),
        "zero_replacement_used": False,
        "median_sugar_share_pct": median_share,
        "ilr_delta_for_median_five_point_contrast": delta,
    }
    return fit, rows, qc


def independent_validation(primary: Fit) -> pd.DataFrame:
    model = LinearRegression(fit_intercept=False)
    model.fit(primary.X, primary.y)
    beta = model.coef_.astype(float)
    residual = primary.y - model.predict(primary.X)
    bread = linalg.pinvh(primary.X.T @ primary.X)
    groups = primary.data["iso3"].to_numpy()
    score_rows = []
    for group in pd.unique(groups):
        mask = groups == group
        score_rows.append(primary.X[mask].T @ residual[mask])
    scores = np.vstack(score_rows)
    meat = scores.T @ scores
    covariance = primary.cr1_factor * bread @ meat @ bread
    se = np.sqrt(np.clip(np.diag(covariance), 0, None))
    j = primary.names.index("sugar5")
    est, std = float(beta[j]), float(se[j])
    cluster_df = primary.groups - 1
    critical = float(stats.t.ppf(0.975, cluster_df))
    return pd.DataFrame([{
        "implementation": "scikit-learn LinearRegression full-dummy fixed effects + independently aggregated CR1 cluster scores",
        "estimate": est,
        "standard_error": std,
        "ci_lower": est - critical * std,
        "ci_upper": est + critical * std,
        "p_value": float(2 * stats.t.sf(abs(est / std), cluster_df)),
        "observations": primary.nobs,
        "economies": primary.groups,
        "cluster_df": cluster_df,
        "coefficient_difference_from_matrix_fit": est - float(primary.beta[j]),
        "se_difference_from_matrix_fit": std - float(primary.standard_error[j]),
        "coefficient_tolerance_pass": bool(abs(est - primary.beta[j]) < 1e-10),
        "se_tolerance_pass": bool(abs(std - primary.standard_error[j]) < 1e-10),
    }])


def wild_cluster_bootstrap_t(primary: Fit, reps: int = 999, seed: int = 20260811) -> pd.DataFrame:
    j = primary.names.index("sugar5")
    x = primary.X[:, j]
    Z = np.delete(primary.X, j, axis=1)
    Q, _ = np.linalg.qr(Z, mode="reduced")
    x_tilde = x - Q @ (Q.T @ x)
    y_tilde = primary.y - Q @ (Q.T @ primary.y)
    xx = float(x_tilde @ x_tilde)
    beta = float(x_tilde @ y_tilde / xx)
    residual = y_tilde - x_tilde * beta
    groups = primary.data["iso3"].to_numpy()
    unique_groups = pd.unique(groups)
    observed_scores = np.array([x_tilde[groups == g] @ residual[groups == g] for g in unique_groups])
    observed_se = math.sqrt(primary.cr1_factor * float(observed_scores @ observed_scores) / (xx * xx))
    observed_t = beta / observed_se

    rng = np.random.default_rng(seed)
    weights = rng.choice(np.array([-1.0, 1.0]), size=(len(unique_groups), reps), replace=True)
    group_codes = pd.Categorical(groups, categories=unique_groups).codes
    y_star = y_tilde[:, None] * weights[group_codes, :]
    y_star_tilde = y_star - Q @ (Q.T @ y_star)
    beta_star = (x_tilde[:, None] * y_star_tilde).sum(axis=0) / xx
    residual_star = y_star_tilde - x_tilde[:, None] * beta_star[None, :]
    cluster_scores = np.zeros((len(unique_groups), reps))
    for gi, group in enumerate(unique_groups):
        mask = groups == group
        cluster_scores[gi] = (x_tilde[mask, None] * residual_star[mask]).sum(axis=0)
    se_star = np.sqrt(primary.cr1_factor * (cluster_scores ** 2).sum(axis=0) / (xx * xx))
    t_star = np.divide(beta_star, se_star, out=np.zeros_like(beta_star), where=se_star > 0)
    p_value = float((1 + np.count_nonzero(np.abs(t_star) >= abs(observed_t))) / (reps + 1))
    return pd.DataFrame([{
        "procedure": "Restricted Rademacher wild-cluster-bootstrap-t",
        "null_hypothesis": "beta_sugar5 = 0",
        "bootstrap_repetitions": reps,
        "seed": seed,
        "observed_estimate": beta,
        "observed_cr1_standard_error": observed_se,
        "observed_t": observed_t,
        "bootstrap_p_value": p_value,
        "bootstrap_t_q025": float(np.quantile(t_star, 0.025)),
        "bootstrap_t_q975": float(np.quantile(t_star, 0.975)),
        "observations": primary.nobs,
        "economies": primary.groups,
    }])


def descriptive_statistics(df: pd.DataFrame) -> pd.DataFrame:
    variables = [
        ("Sugar energy share (%)", "sugar_energy_share_pct"),
        ("Sugar supply (kg/person/year)", "sugar_supply_kg_capita_year"),
        ("Adult obesity (%)", "obesity_adults_pct"),
        ("Premature NCD mortality (%)", "ncd_premature_mortality_probability_pct"),
        ("GDP per capita, PPP ($000)", "gdp_per_capita_ppp_const_2021_intl"),
        ("Disposable-income Gini (points)", "gini_disposable_swiid"),
        ("Working >=49 hours (%)", "working_49plus_pct"),
        ("Urban population (%)", "urban_population_pct"),
        ("Unemployment (%)", "unemployment_pct"),
        ("Internet users (%)", "internet_users_pct"),
        ("Population aged >=65 (%)", "population_age65_plus_pct"),
        ("Health expenditure (% GDP)", "health_expenditure_gdp_pct"),
    ]
    rows = []
    for label, var in variables:
        d = df.dropna(subset=[var]).copy()
        x = d[var].astype(float)
        if var == "gdp_per_capita_ppp_const_2021_intl":
            x = x / 1000
            d = d.assign(_value=x)
            value_col = "_value"
        else:
            value_col = var
        means = d.groupby("iso3")[value_col].mean()
        deviations = d[value_col] - d.groupby("iso3")[value_col].transform("mean")
        rows.append({
            "construct": label,
            "variable": var,
            "observations": len(d),
            "economies": d["iso3"].nunique(),
            "first_year": int(d["year"].min()),
            "last_year": int(d["year"].max()),
            "mean": float(d[value_col].mean()),
            "total_sd": float(d[value_col].std(ddof=1)),
            "between_sd": float(means.std(ddof=1)),
            "within_sd": float(deviations.std(ddof=1)),
            "minimum": float(d[value_col].min()),
            "maximum": float(d[value_col].max()),
        })
    return pd.DataFrame(rows)


def sample_cascade(df: pd.DataFrame) -> pd.DataFrame:
    steps = [
        ("Full economy-year frame", []),
        ("Sugar energy share observed", ["sugar5"]),
        ("Sugar share and adult obesity observed", ["sugar5", "obesity_adults_pct"]),
        ("Primary adjusted complete cases", ["sugar5", "obesity_adults_pct", *CONTROL_VARS]),
        ("Matched temporal complete cases", ["sugar5", "sugar5_lag1", "sugar5_lag3", "sugar5_lead3", "obesity_adults_pct", *CONTROL_VARS]),
        ("Compositional complete cases", ["ilr_sugar_remainder", "obesity_adults_pct", *CONTROL_VARS]),
        ("Inequality interaction complete cases", ["sugar5", "obesity_adults_pct", *CONTROL_VARS, "gini_disposable_swiid"]),
        ("Working-time interaction complete cases", ["sugar5", "obesity_adults_pct", *CONTROL_VARS, "working_49plus_pct"]),
        ("NCD mortality adjusted complete cases", ["sugar5", "ncd_premature_mortality_probability_pct", *CONTROL_VARS]),
        ("Raised-glucose adjusted complete cases", ["sugar5", "raised_fasting_glucose_adults_pct_who", *CONTROL_VARS]),
    ]
    rows = []
    for label, cols in steps:
        d = df.dropna(subset=cols) if cols else df
        rows.append({
            "step": label,
            "observations": len(d),
            "economies": d["iso3"].nunique(),
            "first_year": int(d["year"].min()),
            "last_year": int(d["year"].max()),
            "share_of_full_frame": len(d) / len(df),
        })
    return pd.DataFrame(rows)


def interaction_model(df: pd.DataFrame, context: str, label: str, time_scale: float = 10.0) -> tuple[Fit, dict]:
    # Standardise period means across every economy with at least one observed
    # context value, before restricting to the interaction's complete cases.
    economy_means = df.groupby("iso3")[context].mean().dropna()
    context_mean = float(economy_means.mean())
    context_sd = float(economy_means.std(ddof=1))
    required = ["obesity_adults_pct", "sugar5", *CONTROL_VARS, context, "iso3", "year"]
    d = df.dropna(subset=required).copy().reset_index(drop=True)
    d["context_mean_z"] = d["iso3"].map((economy_means - context_mean) / context_sd)
    d["sugar5_context_interaction"] = d["sugar5"] * d["context_mean_z"]
    xvars = ["sugar5", "sugar5_context_interaction", *CONTROL_VARS]
    context_control = f"{context}_scaled"
    if context not in {"internet_users_pct"}:
        d[context_control] = d[context] / time_scale
        xvars.append(context_control)
    fit = fit_clustered(d, "obesity_adults_pct", xvars, label)
    row = fit.result("sugar5_context_interaction", "interaction", "change in sugar coefficient per 1-SD higher economy-period mean context")
    row.update({"context": label, "context_mean": context_mean, "context_between_sd": context_sd})
    return fit, row


def exact_difference_data(df: pd.DataFrame, years: int, variables: list[str]) -> pd.DataFrame:
    current = df[["iso3", "year", *variables]].copy()
    prior = current.copy()
    prior["year"] = prior["year"] + years
    prior = prior.rename(columns={v: f"{v}_prior" for v in variables})
    merged = current.merge(prior, on=["iso3", "year"], how="left", validate="one_to_one")
    for v in variables:
        merged[f"d{years}_{v}"] = merged[v] - merged[f"{v}_prior"]
    return merged


def fit_difference_model(df: pd.DataFrame, years: int, label: str) -> Fit:
    variables = ["obesity_adults_pct", "sugar5", *CONTROL_VARS]
    d = exact_difference_data(df, years, variables)
    yvar = f"d{years}_obesity_adults_pct"
    xvars = [f"d{years}_sugar5", *[f"d{years}_{v}" for v in CONTROL_VARS]]
    return fit_clustered(d, yvar, xvars, label, economy_fe=False, year_fe=True)


def two_way_cluster_result(primary: Fit) -> dict:
    X, u, d = primary.X, primary.residual, primary.data
    bread = np.linalg.pinv(X.T @ X, hermitian=True)
    n, k = primary.nobs, primary.rank
    country = d["iso3"].to_numpy()
    year = d["year"].to_numpy()
    g1, g2 = len(pd.unique(country)), len(pd.unique(year))
    f1 = (g1 / (g1 - 1)) * ((n - 1) / (n - k))
    f2 = (g2 / (g2 - 1)) * ((n - 1) / (n - k))
    f12 = (n / (n - 1)) * ((n - 1) / (n - k))
    meat = f1 * cluster_meat(X, u, country) + f2 * cluster_meat(X, u, year) - f12 * cluster_meat(X, u, np.arange(n))
    cov = bread @ meat @ bread
    j = primary.names.index("sugar5")
    est, se = float(primary.beta[j]), math.sqrt(max(float(cov[j, j]), 0))
    cluster_df = min(g1, g2) - 1
    critical = float(stats.t.ppf(0.975, cluster_df))
    return {
        "family": "adult_obesity_diagnostic",
        "model": "Two-way clustered SE",
        "term": "sugar5",
        "scale": "per 5-percentage-point higher sugar energy share",
        "estimate": est,
        "standard_error": se,
        "ci_lower": est - critical * se,
        "ci_upper": est + critical * se,
        "p_value": float(2 * stats.t.sf(abs(est / se), cluster_df)),
        "observations": n,
        "economies": g1,
        "cluster_df": cluster_df,
        "first_year": int(d["year"].min()),
        "last_year": int(d["year"].max()),
        "rank": k,
        "df_residual": n - k,
        "cr1_factor": np.nan,
    }


def save_tiff(fig: plt.Figure, path: Path) -> None:
    temporary = path.with_suffix(".unflattened.tif")
    verified = path.with_suffix(".verified.tif")
    fig.savefig(temporary, dpi=350, bbox_inches="tight", pad_inches=0.02, facecolor="white")
    plt.close(fig)
    with Image.open(temporary) as source:
        source.load()
        flattened = Image.new("RGB", source.size, "white")
        if source.mode == "RGBA":
            flattened.paste(source.convert("RGB"), mask=source.getchannel("A"))
        else:
            flattened.paste(source.convert("RGB"))
        max_width, max_height = 2250, 2625
        scale = min(1.0, max_width / flattened.width, max_height / flattened.height)
        if scale < 1.0:
            flattened = flattened.resize(
                (round(flattened.width * scale), round(flattened.height * scale)),
                Image.Resampling.LANCZOS,
            )
        flattened.load()
        flattened.save(verified, format="TIFF", compression="tiff_lzw", dpi=(350, 350))
    with Image.open(verified) as check:
        check.load()
        if check.mode != "RGB":
            raise RuntimeError(f"{verified.name} is not a flattened RGB TIFF")
    os.replace(verified, path)
    temporary.unlink()


def make_fig1(path: Path) -> None:
    fig, ax = plt.subplots(figsize=(7.35, 3.50))
    ax.set_xlim(0, 10)
    ax.set_ylim(0, 5)
    ax.axis("off")
    navy, orange = "#1f4e85", "#d95f02"

    ax.text(
        5, 4.83, "Macro-level constructs represented in the economy–year panel",
        ha="center", va="center", fontsize=10.2, weight="bold",
    )
    top = [
        (0.28, 3.22, 2.45, 1.20, "Food-system\nconditions", "Measured structural\nfood-system indicators"),
        (3.78, 3.22, 2.45, 1.20, "National sugar\navailability", "Per-capita sugar\nsupply/availability"),
        (7.27, 3.22, 2.45, 1.20, "Population metabolic\nand NCD outcomes", "Obesity, diabetes and\ncardiovascular indicators"),
    ]
    for x, y, w, h, title, subtitle in top:
        ax.add_patch(
            FancyBboxPatch(
                (x, y), w, h, boxstyle="round,pad=0.03,rounding_size=0.10",
                edgecolor=navy, facecolor="white", linewidth=1.35,
            )
        )
        ax.text(x + w / 2, y + h * 0.68, title, ha="center", va="center", fontsize=8.8, weight="bold")
        ax.text(x + w / 2, y + h * 0.27, subtitle, ha="center", va="center", fontsize=7.1)
    for x1, x2 in [(2.73, 3.78), (6.23, 7.27)]:
        ax.add_patch(
            FancyArrowPatch(
                (x1, 3.82), (x2, 3.82), arrowstyle="-|>", mutation_scale=16,
                color=navy, linewidth=1.35,
            )
        )

    ax.text(5, 2.74, "Hypothesised mechanisms not directly measured", ha="center", va="center", fontsize=9.4, weight="bold")
    lower = [
        ("Commercial and\nplatform\nexposure", "Marketing, advertising,\ndigital promotion,\nalgorithmic targeting"),
        ("Behavioural\nconditioning", "Cravings, habit\nformation, cue\nreactivity"),
        ("Reward and stress\nregulation", "Neurobiological reward\nresponses and stress\ncoping"),
        ("Dietary behaviour\nand preferences", "Food choices, portion\nsizes, sugar-sweetened\nbeverage intake"),
        ("Metabolic\nadaptation", "Insulin resistance,\ninflammation,\nadiposity"),
    ]
    xs = [0.55, 2.45, 4.35, 6.25, 8.15]
    box_w, box_h, y0 = 1.55, 1.36, 0.76
    for x, (title, subtitle) in zip(xs, lower):
        ax.add_patch(
            FancyBboxPatch(
                (x, y0), box_w, box_h, boxstyle="round,pad=0.025,rounding_size=0.08",
                edgecolor=orange, facecolor="white", linewidth=1.15, linestyle=(0, (4, 3)),
            )
        )
        ax.text(x + box_w / 2, y0 + box_h * 0.70, title, ha="center", va="center", fontsize=6.8, weight="bold", linespacing=0.95)
        ax.text(x + box_w / 2, y0 + box_h * 0.25, subtitle, ha="center", va="center", fontsize=5.7, linespacing=1.0)

    for x1, x2 in zip(xs[:-1], xs[1:]):
        ax.add_patch(
            FancyArrowPatch(
                (x1 + box_w, y0 + box_h / 2), (x2, y0 + box_h / 2),
                arrowstyle="-|>", mutation_scale=9, color=orange, linewidth=1.0,
                linestyle=(0, (4, 3)),
            )
        )
    ax.add_patch(FancyArrowPatch((1.93, 3.22), (1.93, 2.12), arrowstyle="-|>", mutation_scale=14, color=orange, linewidth=1.0, linestyle=(0, (4, 3))))
    ax.plot([5.00, 5.00, 7.03], [3.22, 2.32, 2.32], color=orange, linewidth=1.0, linestyle=(0, (4, 3)))
    ax.add_patch(FancyArrowPatch((7.03, 2.32), (7.03, 2.12), arrowstyle="-|>", mutation_scale=12, color=orange, linewidth=1.0, linestyle=(0, (4, 3))))
    ax.add_patch(FancyArrowPatch((8.88, 2.12), (8.88, 3.22), arrowstyle="-|>", mutation_scale=14, color=orange, linewidth=1.0, linestyle=(0, (4, 3))))

    ax.plot([0.55, 0.55, 9.70, 9.70], [0.52, 0.42, 0.42, 0.52], color=orange, linewidth=0.9)
    ax.text(5.18, 0.13, "Require individual-level, commercial-platform or longitudinal behavioural data", ha="center", va="center", fontsize=6.8)
    save_tiff(fig,path)


def make_fig3(results: pd.DataFrame, path: Path) -> None:
    by_model = results.set_index("model")
    sections = [
        ("Adult obesity — original analytical samples", [
            ("Exposure-only TWFE", "TWFE, exposure only", "#1f4e85", "o", False),
            ("Adjusted TWFE", "Primary TWFE, adjusted", "#1f4e85", "o", True),
            ("Lag 1 year", "One-year lag", "#1f4e85", "o", False),
            ("Lag 3 years", "Three-year lag", "#1f4e85", "o", False),
            ("Future exposure (lead 3)", "Future exposure (three-year lead)", "#1f4e85", "D", False),
            ("Economy-specific trends", "Economy-specific linear trends", "#1f4e85", "o", False),
            ("Annual difference", "Annual first difference", "#1f4e85", "o", False),
            ("Five-year difference", "Five-year difference", "#1f4e85", "o", False),
            ("Population-weighted TWFE", "Population-weighted TWFE", "#1f4e85", "o", False),
            ("Two-way clustered SE", "Two-way clustered SE", "#1f4e85", "o", False),
            ("At least 10 complete years", "At least 10 complete years", "#1f4e85", "o", False),
            ("Microstate/outlier exclusion", "Exclude microstates and exposure outliers", "#1f4e85", "o", False),
            ("Pre-2020 period", "Pre-2020 period", "#1f4e85", "o", False),
        ]),
        ("Common temporal sample (1,228 observations; 156 economies)", [
            ("Matched contemporaneous", "Matched contemporaneous", "#007c83", "o", False),
            ("Matched lag 1 year", "Matched one-year lag", "#007c83", "o", False),
            ("Matched lag 3 years", "Matched three-year lag", "#007c83", "o", False),
            ("Matched future exposure (lead 3)", "Matched future exposure (three-year lead)", "#007c83", "D", False),
        ]),
        ("Compositional sensitivity", [
            ("Two-part ILR balance: median-equivalent 5-point contrast", "Two-part ILR: median-equivalent five-point contrast", "#6a51a3", "^", False),
        ]),
        ("Secondary cardiometabolic outcomes", [
            ("NCD mortality: adjusted TWFE", "Premature NCD mortality: adjusted TWFE", "#d95f02", "s", False),
            ("NCD mortality: lag 3 years", "Premature NCD mortality: three-year lag", "#d95f02", "s", False),
            ("Raised glucose: adjusted TWFE", "Raised fasting glucose: adjusted TWFE", "#d95f02", "s", False),
            ("Raised glucose: lag 3 years", "Raised fasting glucose: three-year lag", "#d95f02", "s", False),
        ]),
    ]
    display_rows = []
    for heading, items in sections:
        display_rows.append(("heading", heading, None))
        for item in items:
            display_rows.append(("estimate", item[1], item))

    fig, ax = plt.subplots(figsize=(7.35, 7.15))
    fig.subplots_adjust(left=0.43, right=0.74, top=0.965, bottom=0.09)
    n = len(display_rows)
    y_positions = np.arange(n)[::-1]
    ax.set_ylim(-0.8, n - 0.2)
    ax.set_xlim(-1.45, 0.60)
    ax.axvline(0, color="#666666", linewidth=0.85, zorder=0)

    for y, (kind, label, payload) in zip(y_positions, display_rows):
        if kind == "heading":
            ax.text(-0.04, y, label, transform=ax.get_yaxis_transform(), ha="right", va="center",
                    fontsize=7.45, weight="bold", color="#111111", clip_on=False)
            continue
        model_name, _, colour, marker, primary = payload
        row = by_model.loc[model_name]
        ax.errorbar(
            row["estimate"], y,
            xerr=[[row["estimate"] - row["ci_lower"]], [row["ci_upper"] - row["estimate"]]],
            fmt=marker, color=colour, ecolor=colour, markerfacecolor="white" if marker == "D" else colour,
            markeredgewidth=1.0, markersize=4.2 if not primary else 5.1, linewidth=1.15, capsize=0, zorder=3,
        )
        ax.text(-0.04, y, label, transform=ax.get_yaxis_transform(), ha="right", va="center",
                fontsize=7.25, weight="bold" if primary else "normal", clip_on=False)
        estimate_text = f"{row['estimate']:.2f} ({row['ci_lower']:.2f} to {row['ci_upper']:.2f})".replace("-", "−")
        ax.text(1.07, y, estimate_text, transform=ax.get_yaxis_transform(), ha="left", va="center",
                fontsize=7.25, clip_on=False)

    ax.text(-0.04, n - 0.02, "Specification", transform=ax.get_yaxis_transform(), ha="right", va="bottom",
            fontsize=7.7, weight="bold", clip_on=False)
    ax.text(1.07, n - 0.02, "Estimate (95% CI)", transform=ax.get_yaxis_transform(), ha="left", va="bottom",
            fontsize=7.7, weight="bold", clip_on=False)
    ax.set_yticks([])
    ax.set_xlabel("Outcome difference (percentage points) per comparable five-point exposure contrast", fontsize=7.4)
    ax.tick_params(axis="x", labelsize=7.2, length=3)
    ax.spines[["top", "right", "left"]].set_visible(False)
    ax.grid(axis="x", color="#e6e6e6", linewidth=0.45, zorder=0)
    save_tiff(fig,path)


def make_fig4(interactions: pd.DataFrame, path: Path) -> None:
    fig, ax = plt.subplots(figsize=(7.35, 3.20))
    fig.subplots_adjust(left=0.31, right=0.65, top=0.82, bottom=0.20)
    blue, grey = "#1f77b4", "#666a70"
    descriptions = {
        "Income inequality": "Disposable-income Gini",
        "Long-hours work": "Employed persons working ≥49 h/week",
        "Digital access": "Individuals using the Internet",
    }
    y = np.arange(len(interactions))[::-1]
    for yi, (_, row) in zip(y, interactions.iterrows()):
        ax.errorbar(
            row.estimate, yi,
            xerr=[[row.estimate - row.ci_lower], [row.ci_upper - row.estimate]],
            fmt="o", color=blue, ecolor="#202020", markeredgecolor="white",
            markeredgewidth=0.5, markersize=5.2, linewidth=1.25, capsize=3,
        )
    ax.axvline(0, color="#666666", linestyle=(0, (4, 3)), linewidth=1.0)
    ax.set_yticks([])

    for yi, (_, row) in zip(y, interactions.iterrows()):
        ax.text(-0.08, yi + 0.10, row.context, transform=ax.get_yaxis_transform(), ha="right", va="center", fontsize=8.0)
        ax.text(-0.08, yi - 0.17, descriptions[row.context], transform=ax.get_yaxis_transform(), ha="right", va="center", fontsize=6.9, color=grey)
        estimate = f"{row.estimate:.2f} ({row.ci_lower:.2f} to {row.ci_upper:.2f})".replace("-", "−")
        detail = f"$P$ = {row.p_value:.3f}; q = {row.q_value_bh:.3f}  ·  {int(row.observations):,} / {int(row.economies)}"
        ax.text(1.10, yi + 0.10, estimate, transform=ax.get_yaxis_transform(), ha="left", va="center", fontsize=7.9)
        ax.text(1.10, yi - 0.17, detail, transform=ax.get_yaxis_transform(), ha="left", va="center", fontsize=6.9, color=grey)
    for yi in [0.5, 1.5]:
        ax.axhline(yi, color="#dddddd", linewidth=0.65, xmin=-0.82, xmax=2.38, clip_on=False)

    fig.text(0.045, 0.95, "Cross-level modification by structural context", ha="left", va="center", fontsize=9.7, weight="bold")
    ax.text(-0.08, 1.03, "Modifier", transform=ax.transAxes, ha="right", va="bottom", fontsize=7.7, weight="bold", color=grey)
    ax.text(0.50, 1.03, "Interaction estimate (95% CI)", transform=ax.transAxes, ha="center", va="bottom", fontsize=7.3, weight="bold", color=grey)
    ax.text(1.10, 1.03, "Δβ (95% CI)\nP; q  ·  observations / economies", transform=ax.transAxes, ha="left", va="bottom", fontsize=7.4, weight="bold", color=grey)
    ax.set_xlabel("Change in sugar–obesity coefficient, Δβ\nper 1-SD higher economy-period mean context", fontsize=7.7)
    ax.set_xlim(-1.25, 0.75)
    ax.set_ylim(-0.35, 2.35)
    ax.set_xticks([-1.0, -0.5, 0.0, 0.5])
    ax.tick_params(axis="x", labelsize=7.3, length=3)
    ax.spines[["top", "right", "left"]].set_visible(False)
    save_tiff(fig, path)


def make_fig2(df: pd.DataFrame, path: Path, figure_data_path: Path) -> None:
    annual = []
    for year, d in df.groupby("year"):
        for var in ["sugar_energy_share_pct","obesity_adults_pct"]:
            q=d.dropna(subset=[var,"population_total"])
            annual.append({"year":year,"variable":var,"population_weighted_mean":float(np.average(q[var],weights=q.population_total))})
    annual_df=pd.DataFrame(annual)
    joint=df.dropna(subset=["sugar_energy_share_pct","obesity_adults_pct"])
    means=joint.groupby(["iso3","country_name","region"],as_index=False)[["sugar_energy_share_pct","obesity_adults_pct"]].mean()
    starts=joint[joint.year.eq(2010)][["iso3","sugar_energy_share_pct","obesity_adults_pct"]].rename(columns={"sugar_energy_share_pct":"sugar_start","obesity_adults_pct":"obesity_start"})
    ends=joint.sort_values("year").groupby("iso3",as_index=False).tail(1)[["iso3","year","sugar_energy_share_pct","obesity_adults_pct"]]
    changes=starts.merge(ends,on="iso3",how="inner")
    changes["sugar_change"]=changes.sugar_energy_share_pct-changes.sugar_start
    changes["obesity_change"]=changes.obesity_adults_pct-changes.obesity_start
    pd.concat([
        annual_df.assign(panel="annual"),
        means.rename(columns={"sugar_energy_share_pct":"x","obesity_adults_pct":"y"}).assign(panel="between"),
        changes.rename(columns={"sugar_change":"x","obesity_change":"y"}).assign(panel="change"),
    ],ignore_index=True,sort=False).to_csv(figure_data_path,index=False)
    fig, axs = plt.subplots(2, 2, figsize=(7.35, 5.82))
    blue = "#1f77b4"
    for label, ax in zip("abcd", axs.flat):
        ax.text(0.01, 1.04, label, transform=ax.transAxes, fontsize=9.5, weight="bold")
        ax.tick_params(labelsize=7.5, length=3)
        ax.spines[["top", "right"]].set_visible(False)

    trend_specs = [
        (axs[0, 0], "sugar_energy_share_pct", "Population-weighted sugar and sweeteners\nshare of dietary energy", "Sugar and sweeteners share\nof dietary energy (%)"),
        (axs[0, 1], "obesity_adults_pct", "Population-weighted age-standardised\nadult obesity prevalence", "Age-standardised adult\nobesity prevalence (%)"),
    ]
    for ax, variable, title, ylabel in trend_specs:
        panel = annual_df[annual_df.variable.eq(variable)]
        ax.plot(panel.year, panel.population_weighted_mean, "o-", color=blue, markersize=3.2, linewidth=1.15)
        ax.set_title(title, fontsize=8.8, weight="bold", pad=5)
        ax.set_xlabel("Year", fontsize=7.8)
        ax.set_ylabel(ylabel, fontsize=7.8)

    region_style = {
        "East Asia & Pacific": ("#1f77b4", "o"),
        "Europe & Central Asia": ("#ff7f0e", "s"),
        "Latin America & Caribbean": ("#2ca02c", "^"),
        "Middle East & North Africa": ("#d62728", "D"),
        "North America": ("#9467bd", "P"),
        "South Asia": ("#8c564b", "v"),
        "Sub-Saharan Africa": ("#e377c2", "X"),
    }
    for region, group in means.groupby("region"):
        colour, marker = region_style.get(region, ("#7f7f7f", "o"))
        axs[1, 0].scatter(
            group.sugar_energy_share_pct, group.obesity_adults_pct,
            s=20, label=region, color=colour, marker=marker,
            edgecolor="#333333", linewidth=0.35, alpha=0.85,
        )
    slope, intercept = np.polyfit(means.sugar_energy_share_pct, means.obesity_adults_pct, 1)
    xx = np.linspace(means.sugar_energy_share_pct.min(), means.sugar_energy_share_pct.max(), 100)
    axs[1, 0].plot(xx, intercept + slope * xx, color="#111111", linewidth=1.0)
    rho, pv = stats.spearmanr(means.sugar_energy_share_pct, means.obesity_adults_pct)
    axs[1, 0].text(
        0.03, 0.95, f"Spearman $\\rho$ = {rho:.2f}\n$P$ < 0.001; $n$ = {len(means)}",
        transform=axs[1, 0].transAxes, va="top", fontsize=7.2,
    )
    axs[1, 0].set_xlabel("Mean sugar and sweeteners share\nof dietary energy (%)", fontsize=7.8)
    axs[1, 0].set_ylabel("Mean age-standardised adult\nobesity prevalence (%)", fontsize=7.8)
    axs[1, 0].set_title("Between-economy levels\nEconomy-period means by World Bank region", fontsize=8.5, weight="bold", pad=5)

    axs[1, 1].scatter(
        changes.sugar_change, changes.obesity_change, s=18,
        facecolors="white", edgecolors="#111111", linewidth=0.7,
    )
    slope2, intercept2 = np.polyfit(changes.sugar_change, changes.obesity_change, 1)
    xx2 = np.linspace(changes.sugar_change.min(), changes.sugar_change.max(), 100)
    axs[1, 1].plot(xx2, intercept2 + slope2 * xx2, color="#111111", linewidth=1.0)
    rho2, pv2 = stats.spearmanr(changes.sugar_change, changes.obesity_change)
    axs[1, 1].text(
        0.97, 0.95, f"Spearman $\\rho$ = {rho2:.2f}\n$P$ = {pv2:.4f}; $n$ = {len(changes)}",
        transform=axs[1, 1].transAxes, ha="right", va="top", fontsize=7.2,
    )
    axs[1, 1].axhline(0, color="#888888", linestyle=(0, (3, 2)), linewidth=0.7)
    axs[1, 1].axvline(0, color="#888888", linestyle=(0, (3, 2)), linewidth=0.7)
    axs[1, 1].set_xlabel("Change in sugar and sweeteners share\nof dietary energy (percentage points)", fontsize=7.8)
    axs[1, 1].set_ylabel("Change in age-standardised adult obesity\nprevalence (percentage points)", fontsize=7.8)
    axs[1, 1].set_title("Within-economy changes\nFrom 2010 to last jointly observed year", fontsize=8.5, weight="bold", pad=5)

    handles, labels = axs[1, 0].get_legend_handles_labels()
    fig.legend(handles, labels, loc="lower center", bbox_to_anchor=(0.5, 0.012), ncol=4, fontsize=6.8, frameon=False, handletextpad=0.5, columnspacing=1.2)
    fig.subplots_adjust(left=0.09, right=0.99, top=0.95, bottom=0.22, hspace=0.55, wspace=0.36)
    save_tiff(fig, path)


def main() -> None:
    parser = argparse.ArgumentParser()
    parser.add_argument("--data", type=Path, default=Path("S1_Data.xlsx"))
    parser.add_argument("--output", type=Path, default=Path("outputs"))
    parser.add_argument("--bootstrap-reps", type=int, default=999)
    args = parser.parse_args()
    args.output.mkdir(parents=True, exist_ok=True)
    df = read_masterfile(args.data)

    model_rows: list[dict] = []
    _, mundlak_rows = mundlak_model(df)
    model_rows.extend(mundlak_rows)

    exposure_only = fit_clustered(df,"obesity_adults_pct",["sugar5"],"Exposure-only TWFE")
    primary = fit_clustered(df,"obesity_adults_pct",["sugar5",*CONTROL_VARS],"Adjusted TWFE")
    lag1 = fit_clustered(df,"obesity_adults_pct",["sugar5_lag1",*CONTROL_VARS],"Lag 1 year")
    lag3 = fit_clustered(df,"obesity_adults_pct",["sugar5_lag3",*CONTROL_VARS],"Lag 3 years")
    lead3 = fit_clustered(df,"obesity_adults_pct",["sugar5_lead3",*CONTROL_VARS],"Future exposure (lead 3)")
    trends = fit_clustered(df,"obesity_adults_pct",["sugar5",*CONTROL_VARS],"Economy-specific trends",economy_trends=True)
    annual_diff = fit_difference_model(df,1,"Annual difference")
    five_diff = fit_difference_model(df,5,"Five-year difference")
    pop_weighted = fit_clustered(df,"obesity_adults_pct",["sugar5",*CONTROL_VARS],"Population-weighted TWFE",weights="population_total")
    for fit,term in [(exposure_only,"sugar5"),(primary,"sugar5"),(lag1,"sugar5_lag1"),(lag3,"sugar5_lag3"),(lead3,"sugar5_lead3"),(trends,"sugar5"),(annual_diff,"d1_sugar5"),(five_diff,"d5_sugar5"),(pop_weighted,"sugar5")]:
        model_rows.append(fit.result(term,"adult_obesity_diagnostic","per 5-percentage-point higher sugar energy share"))
    model_rows.append(two_way_cluster_result(primary))

    complete_counts=primary.data.groupby('iso3').size()
    at_least10=df[df.iso3.isin(complete_counts[complete_counts>=10].index)]
    well_observed=fit_clustered(at_least10,"obesity_adults_pct",["sugar5",*CONTROL_VARS],"At least 10 complete years")
    # The legacy diagnostic used the 99th percentile of the source mass measure;
    # after the population restriction, no additional complete-case row exceeds
    # this threshold, but the rule is retained for exact reproduction.
    primary_mass_q99=float(primary.data.sugar_supply_kg_capita_year.quantile(.99))
    restricted=df[(df.population_total>=1_000_000)&(df.sugar_supply_kg_capita_year<=primary_mass_q99)]
    outlier=fit_clustered(restricted,"obesity_adults_pct",["sugar5",*CONTROL_VARS],"Microstate/outlier exclusion")
    pre2020=fit_clustered(df[df.year<=2019],"obesity_adults_pct",["sugar5",*CONTROL_VARS],"Pre-2020 period")
    kg=fit_clustered(df,"obesity_adults_pct",["sugar_kg10",*CONTROL_VARS],"Sugar supply mass (per 10 kg)")
    kcal=fit_clustered(df,"obesity_adults_pct",["sugar_kcal100",*CONTROL_VARS],"Sugar energy (per 100 kcal)")
    for fit,term,scale in [(well_observed,"sugar5","per 5-percentage-point higher sugar energy share"),(outlier,"sugar5","per 5-percentage-point higher sugar energy share"),(pre2020,"sugar5","per 5-percentage-point higher sugar energy share"),(kg,"sugar_kg10","per 10 kg/person/year"),(kcal,"sugar_kcal100","per 100 kcal/person/day")]:
        model_rows.append(fit.result(term,"adult_obesity_diagnostic",scale))

    secondary=[]
    for outcome,label in [("ncd_premature_mortality_probability_pct","NCD mortality: adjusted TWFE"),("raised_fasting_glucose_adults_pct_who","Raised glucose: adjusted TWFE")]:
        f=fit_clustered(df,outcome,["sugar5",*CONTROL_VARS],label); r=f.result("sugar5","secondary_outcome","per 5-percentage-point higher sugar energy share"); secondary.append(r); model_rows.append(r)
        f3=fit_clustered(df,outcome,["sugar5_lag3",*CONTROL_VARS],label.replace("adjusted TWFE","lag 3 years")); r3=f3.result("sugar5_lag3","secondary_outcome","per 5-percentage-point higher sugar energy share"); secondary.append(r3); model_rows.append(r3)

    matched_fits, matched_rows, contrasts = matched_temporal(df)
    matched_exposures = matched_fits[0].data[["sugar5", "sugar5_lag1", "sugar5_lag3", "sugar5_lead3"]]
    matched_correlations = matched_exposures.corr(method="pearson")
    matched_correlations.index.name = "exposure"
    model_rows.extend(matched_rows.to_dict("records"))
    comp_fit, comp_rows, comp_qc = compositional_model(df)
    model_rows.extend(comp_rows.to_dict("records"))

    interactions=[]
    for context,label,scale in [("gini_disposable_swiid","Income inequality",10.0),("working_49plus_pct","Long-hours work",10.0),("internet_users_pct","Digital access",10.0)]:
        _,row=interaction_model(df,context,label,scale); interactions.append(row)
    interactions_df=pd.DataFrame(interactions)
    interactions_df["q_value_bh"] = benjamini_hochberg(interactions_df["p_value"])

    independent=independent_validation(primary)
    wild=wild_cluster_bootstrap_t(primary,reps=args.bootstrap_reps)
    descriptions=descriptive_statistics(df)
    cascade=sample_cascade(df)
    results=pd.DataFrame(model_rows)

    # Residual AR(1) diagnostic on exact adjacent years.
    resid=primary.data[["iso3","year"]].copy(); resid["residual"]=primary.residual
    prior=resid.rename(columns={"residual":"residual_lag1"}).copy(); prior["year"]+=1
    resid_pair=resid.merge(prior,on=["iso3","year"],how="inner")
    ar1=float(np.corrcoef(resid_pair.residual,resid_pair.residual_lag1)[0,1])

    outputs={
        "model_results.csv":results,
        "matched_temporal_results.csv":matched_rows,
        "matched_temporal_contrasts.csv":contrasts,
        "matched_exposure_correlations.csv":matched_correlations.reset_index(),
        "compositional_results.csv":comp_rows,
        "independent_validation.csv":independent,
        "wild_cluster_bootstrap.csv":wild,
        "descriptive_statistics.csv":descriptions,
        "sample_cascade.csv":cascade,
        "interaction_results.csv":interactions_df,
        "secondary_outcomes.csv":pd.DataFrame(secondary),
    }
    for filename,frame in outputs.items(): frame.to_csv(args.output/filename,index=False)
    export_documentation_sheets(args.data, args.output)

    make_fig1(args.output/"Fig1.tif")
    make_fig2(df,args.output/"Fig2.tif",args.output/"figure2_data.csv")
    fig3_order=[
        "Exposure-only TWFE","Adjusted TWFE","Lag 1 year","Lag 3 years","Future exposure (lead 3)","Economy-specific trends","Annual difference","Five-year difference","Population-weighted TWFE","Two-way clustered SE","At least 10 complete years","Microstate/outlier exclusion","Pre-2020 period","Matched contemporaneous","Matched lag 1 year","Matched lag 3 years","Matched future exposure (lead 3)","Two-part ILR balance: median-equivalent 5-point contrast","NCD mortality: adjusted TWFE","NCD mortality: lag 3 years","Raised glucose: adjusted TWFE","Raised glucose: lag 3 years"
    ]
    fig3=results[results.model.isin(fig3_order)].copy(); fig3["_order"]=fig3.model.map({x:i for i,x in enumerate(fig3_order)}); fig3=fig3.sort_values("_order"); fig3.to_csv(args.output/"figure3_data.csv",index=False); make_fig3(fig3,args.output/"Fig3.tif")
    interactions_df.to_csv(args.output/"figure4_data.csv",index=False); make_fig4(interactions_df,args.output/"Fig4.tif")

    summary={
        "data_file":str(args.data.name),
        "master_rows":len(df),
        "master_economies":int(df.iso3.nunique()),
        "primary":primary.result("sugar5"),
        "matched_sample":{"observations":int(matched_rows.observations.iloc[0]),"economies":int(matched_rows.economies.iloc[0]),"first_year":int(matched_rows.first_year.iloc[0]),"last_year":int(matched_rows.last_year.iloc[0])},
        "matched_temporal_conclusion":"Temporal instability survives common-sample matching: contemporaneous and lagged coefficients are near zero, the three-year future-exposure coefficient remains negative, and the future-minus-contemporaneous contrast is -0.578570 (P=0.002213).",
        "compositional_qc":comp_qc,
        "independent_validation_pass":bool(independent.coefficient_tolerance_pass.iloc[0] and independent.se_tolerance_pass.iloc[0]),
        "wild_cluster_bootstrap_p":float(wild.bootstrap_p_value.iloc[0]),
        "residual_ar1_correlation":ar1,
        "software":{"python":os.sys.version.split()[0],"pandas":pd.__version__,"numpy":np.__version__,"scipy":scipy.__version__,"scikit_learn":"1.8.0","matplotlib":matplotlib.__version__},
    }
    (args.output/"analysis_summary.json").write_text(json.dumps(summary,indent=2),encoding="utf-8")
    write_manifest(args.output)
    print(json.dumps(summary,indent=2))


if __name__ == "__main__":
    main()
```
