## Supplementary material for "Inferential instability of national sugar and sweetener availability as an indicator of adult obesity trajectories: A global within–between panel audit": S2 Code: README.md

# S2 Code — Reproducibility instructions

This bundle reproduces the reported economy-year panel analyses and executes the added matched-sample temporal, compositional, independent-validation and wild-cluster-bootstrap checks.

## Required input

Place `S1_Data.xlsx` in the working directory. The script reads the `Master_CountryYear` sheet and, for portable documentation, exports the Coverage, Codebook, Provenance, Country_Crosswalk and Merge_QC sheets to CSV.

## Environment

Python 3.12 was used for the verified run. Install the locked dependencies with:

```bash
python -m pip install -r requirements.txt
```

## Execute

```bash
python analysis.py --data S1_Data.xlsx --output outputs --bootstrap-reps 999
```

The bootstrap seed is fixed at `20260811`. The run writes machine-readable estimates, diagnostics, metadata snapshots, four TIFF figures, `analysis_summary.json` and a SHA-256 manifest. All statistical tests are two-sided. Fixed-effects standard errors use an economy-clustered CR1 covariance unless otherwise stated.

Figures are regenerated at 350 dpi with an Arial-family sans-serif hierarchy. In the verified environment, Matplotlib uses Nimbus Sans, the installed metrically compatible fallback. The TIFF outputs are flattened RGB files with LZW compression.

## Main added checks

- Common temporal sample: current, one-year-lagged, three-year-lagged and three-year-future exposure models use exactly the same outcome-year rows and covariates. Cross-model Wald contrasts use a stacked economy-clustered covariance.
- Composition: a two-part isometric log-ratio balance, `sqrt(1/2) × ln(sugar kcal/non-sugar kcal)`, is estimated without zero replacement because both components are strictly positive in the analytical sample.
- Independent validation: the full-dummy fixed-effects coefficient is re-estimated with scikit-learn and its CR1 clustered covariance is independently reconstructed from cluster score vectors.
- Bootstrap: a restricted Rademacher wild-cluster-bootstrap-t test checks the principal sugar coefficient with 999 repetitions.

## Output interpretation

The manuscript reports rounded values. The CSV files preserve full numerical precision. `analysis_manifest.csv` records the size and SHA-256 checksum of every generated output.
